# Racial Inequalities in Alcohol Use Disorder Diagnosis in a Sample of 700,000 Veterans

**DOI:** 10.1101/2021.07.14.21256113

**Authors:** Rachel Vickers-Smith, Amy C. Justice, William C. Becker, Christopher T. Rentsch, Brenda Curtis, Anita Fernander, Emily E. Hartwell, Eseosa T. Ighodaro, Rachel L. Kember, Janet Tate, Henry R. Kranzler

## Abstract

**Background:** Studies show that Black and Hispanic Veterans have a higher prevalence of alcohol use disorder (AUD) than White Veterans. We examined whether the relationship between self-reported race/ethnicity and AUD diagnosis varies by self-reported alcohol consumption.

**Methods:** The sample included 700,013 Black, Hispanic, and White Veterans enrolled in the Million Veteran Program cohort. Alcohol consumption was defined as an individual’s maximum score on the Alcohol Use Disorders Identification Test–Consumption (AUDIT-C) questionnaire, a screen for hazardous or harmful drinking. The primary outcome, AUD, was defined by the presence of ICD-9/10 codes in the electronic health record. We used logistic regression with interactions to assess the association between race/ethnicity and AUD by maximum AUDIT-C score.

**Results:** Black and Hispanic Veterans were more likely to have an AUD diagnosis than White Veterans despite similar levels of alcohol consumption. The difference was greatest between Black and White men. At all but the lowest and highest levels of alcohol consumption, Black men had 24%-111% greater odds of an AUD diagnosis. The association between race/ethnicity and AUD diagnosis remained after adjustment for alcohol consumption, alcohol-related disorders, and other potential confounders.

**Conclusions:** The large discrepancy in AUD diagnosis across groups despite a similar distribution of alcohol consumption measures suggests that Veterans are differentially assigned an AUD diagnosis by race/ethnicity. Efforts are needed to examine the causes of the observed differences and to implement changes, such as structured diagnostic methods, to address a likely contributor to racial differences (i.e., bias) in AUD diagnosis.

## Introduction

Clinical diagnosis is a foundation of decision making and treatment.^1^ Clinical “labels,” can produce lasting stigma and when inappropriate can cause lasting damage.^2,3^ Further, misdiagnosis (i.e., over or under diagnosis, misidentification) can result in ineffective treatment, inaccurate prognostic assessments, poor outcomes, and mistrust in the healthcare system.^4,5^ For diagnosing behavior-based conditions, factors that can influence diagnosis include multiple medical conditions, varying symptom presentation, provider’s level of education and experience, the patient’s willingness to disclose symptomatology, cultural factors, and the application of standard criteria or assessments.^6^ The race or ethnicity of a patient and patient-provider racial/ethnic discordance, can also influence diagnostic decisions through implicit bias,^4,7,8^ i.e., healthcare providers’ unconscious prejudices or stereotypes that impacts clinical judgment or treatment.^9,10^

Studies in the Veterans Health Administration of the Department of Veterans Affairs (VA) have shown that, compared to White Veterans, Black and Hispanic Veterans have a higher rate of clinically recognized alcohol use disorder (AUD)^11^ and Black Veterans are more likely to be identified as needing an intervention^12,13^ and to receive psychosocial interventions,^12,14^ but less likely to receive pharmacotherapy for AUD.^15^ One potential explanation for the observed disparities is that the groups have different alcohol consumption patterns.^16^

Here, we examine the contribution of self-reported alcohol consumption to the likelihood of receiving an AUD diagnosis among Black, Hispanic, and White Veterans. To facilitate the identification of individuals with at-risk or harmful drinking, beginning in 2008 the VA has routinely screened primary care patients using the AUDIT-C^17,18^ (the first three items of the 10-item Alcohol Use Disorders Identification Test^19^). We examined the association of AUDIT-C scores with AUD diagnosis codes across the three racial/ethnic groups in a national cohort of more than 700,000 Veterans. Specifically, we evaluated: (1) the relationship between race/ethnicity and AUD diagnosis across similar levels of self-reported alcohol consumption and (2) sociodemographic and clinical correlates of an AUD diagnosis.

## Methods

Due to space constraints, abbreviated methods are described below; detailed methods can be found in the Supplementary Appendix.

### Study Sample

The study sample included 700,013 participants in the Million Veteran Program (MVP), an ongoing longitudinal cohort of US Veterans.^20^

### Measures

#### Race/ethnicity

Race/ethnicity was self-reported as non-Hispanic White (White), non-Hispanic Black (Black), and Hispanic. Participants who self-identified as Hispanic were so classified irrespective of their race, consistent with the U.S. Census Bureau.^22^

#### Self-reported alcohol consumption

The AUDIT-C is a valid, reliable screening instrument that is routinely used to identify individuals with hazardous or harmful drinking.^17,18^ Maximal reported alcohol consumption was determined using the highest AUDIT-C recorded in the EHR. We also examined the association between the maximum score on AUDIT-C item 3 (binge drinking frequency) and AUD diagnosis by race/ethnicity.

#### Demographics and clinical diagnoses

Sex and age at enrollment were extracted from both questionnaires and EHR data.^21^ Clinical diagnoses, including AUD, alcohol-related medical disorders (i.e., cirrhosis, neuropathy, cardiomyopathy, gastritis, fatty liver disease, hepatitis, and liver damage), drug use disorder, and mental disorder, required the presence of one inpatient or two outpatient ICD-9/10 diagnostic codes in the VA EHR.

### Data Analysis

All analyses were stratified by sex to account for sex-related differences.^23^ Descriptive statistics by racial/ethnic group were computed with means, standard deviations, and frequencies. We compared characteristics across race/ethnicity using ANOVA with Tukey’s post-hoc tests, chi-square, and Fisher’s exact tests. The correlation between maximum AUDIT-C score and AUD diagnosis was computed using Spearman’s rho, and between binge drinking frequency and AUD diagnosis with a chi-square test. We used logistic regression to identify the factors associated with documented AUD diagnosis and assessed the interaction between race/ethnicity and maximum AUDIT-C score by creating a composite variable of the two factors. We probed significant interactions with stratified logistic regression models. Sensitivity analyses with age-adjusted mean AUDIT-C score assessed the robustness of estimates using maximum AUDIT-C scores. For all models, C-statistics served to assess discrimination. All analyses were performed using SAS 9.2 (Cary, NC), with an alpha of 0.05 to denote statistical significance.

## Results

Of Veterans included in this study, 91% were men (n=638,205) and 9% were women (n=61,808).

### Men

#### Sample Characteristics

The racial/ethnic distribution was White (74%), Black (19%), and Hispanic (7%; Table 1). The modal maximal AUDIT-C score range was 1-3 across the three groups. Black male Veterans (BMV) had more AUDIT-C assessments (mean=9.5; SD=4.6) than Hispanic (HMV; mean=8.3; SD=4.3) or White (WMV; mean=8.8; SD=4.3) male Veterans, and higher frequency of an AUD diagnosis (33%, 25%, and 18%, respectively) and a drug use disorder diagnosis (29%, 16%, and 11%, respectively) than HMV or WMV. Although statistically significant, differences in the prevalence of alcohol-related diagnoses across racial/ethnic groups were small. WMV had a lower frequency of a mental disorder diagnosis (47%) than BMV (58%) or HMV (61%).

**Table 1.** Demographic and clinical characteristics of the sample, by sex and race/ethnicity (n=700,013).

|  | Black | Hispanic | White |  |
| --- | --- | --- | --- | --- |
| **Men (n=638,205)** | 18.6% (n=118,600) | 7.1% (n=45,330) | 74.3% (n=474,275) | *P* |
| Age (y) – mean (SD)* | 59.2 (11.7)<sup>a,b</sup> | 56.0 (15.4)<sup>a,c</sup> | 64.7 (13.2)<sup>b,c</sup> | <0.0001 |
| *Alcohol-Related Characteristics* |  |  |  |  |
| Highest AUDIT-C |  |  |  | <0.0001 |
| 0 | 21.7% (25,733)<sup>a</sup> | 16.9% (7,637)<sup>a,c</sup> | 21.7% (103,113)<sup>c</sup> |  |
| 1-3 | 39.1% (46,371)<sup>a,b</sup> | 42.1% (19,090)<sup>a,c</sup> | 40.9% (193,974)<sup>b,c</sup> |  |
| 4-7 | 23.6% (28,004)<sup>a,b</sup> | 25.4% (11,531)<sup>a,c</sup> | 26.7% (126,710)<sup>b,c</sup> |  |
| 8+ | 15.6% (18,492)<sup>b</sup> | 15.6% (7,072)<sup>c</sup> | 10.7% (50,505)<sup>b,c</sup> |  |
| Number of AUDIT-C assessments – mean (SD) | 9.5 (4.6)<sup>a,b</sup> | 8.3 (4.3)<sup>a,c</sup> | 8.8 (4.3)<sup>b,c</sup> | <0.0001 |
| AUD | 33.4% (39,560)<sup>a,b</sup> | 24.8% (11,227)<sup>a,c</sup> | 18.0% (85,398)<sup>b,c</sup> | <0.0001 |
| Cirrhosis** | 1.5% (1,825)<sup>a,b</sup> | 2.3% (1,025)<sup>a,c</sup> | 1.4% (6,404)<sup>b,c</sup> | <0.0001 |
| Neuropathy** | 0.3% (305)<sup>a</sup> | 0.1% (48)<sup>a,c</sup> | 0.3% (1,188)<sup>c</sup> | <0.0001 |
| Cardiomyopathy** | 0.3% (395)<sup>a,b</sup> | 0.1% (54)<sup>a</sup> | 0.2% (726)<sup>b</sup> | <0.0001 |
| Gastritis** | 0.4% (457)<sup>a,b</sup> | 0.3% (114)<sup>a</sup> | 0.2% (1,111)<sup>b</sup> | <0.0001 |
| Fatty liver disease** | 0.5% (619)<sup>a,b</sup> | 0.7% (310)<sup>a,c</sup> | 0.5% (2,209)<sup>b,c</sup> | <0.0001 |
| Hepatitis** | 0.7% (842)<sup>b</sup> | 0.7% (296)<sup>c</sup> | 0.5% (2,530)<sup>b,c</sup> | <0.0001 |
| Liver damage** | 0.4% (517)<sup>b</sup> | 0.5% (226)<sup>c</sup> | 0.3% (1,495)<sup>b,c</sup> | <0.0001 |
| *Other Clinical ICD-9/10 Diagnoses* |  |  |  |  |
| Drug abuse/dependence | 29.0% (34,443)<sup>a,b</sup> | 16.2% (7,347)<sup>a,c</sup> | 10.8% (51,143)<sup>b,c</sup> | <0.0001 |
| AUD + drug abuse/dependence | 22.9% (27,191)<sup>a,b</sup> | 12.0% (5,439)<sup>a,c</sup> | 7.5% (35,347)<sup>b,c</sup> | <0.0001 |
| Mental disorder | 57.6% (68,290)<sup>a,b</sup> | 60.6% (27,455)<sup>a,c</sup> | 46.5% (220,537)<sup>b,c</sup> | <0.0001 |
| AUD + mental disorder | 26.2% (31,080)<sup>a,b</sup> | 20.8% (9,439)<sup>a,c</sup> | 13.8% (65,388)<sup>b,c</sup> | <0.0001 |
|  | Black | Hispanic | White | *P* |
| **Women (n=61,808)** | 30.0% (n=18,460) | 8.2% (n=5,092) | 61.9% (n=38,256) |  |
| Age (y) – mean (SD)* | 49.2 (11.3)<sup>a,b</sup> | 43.6 (13.1)<sup>a,c</sup> | 52.6 (13.9)<sup>b,c</sup> | <0.0001 |

| *Alcohol-Related Characteristics* |  |  |  |  |
| --- | --- | --- | --- | --- |
| Highest AUDIT-C |  |  |  | <0.0001 |
| 0 | 19.9% (3,664)<sup>a,b</sup> | 13.6% (694)<sup>a,c</sup> | 17.5% (6,692)<sup>b,c</sup> |  |
| 1-2 | 58.3% (10,762)<sup>b</sup> | 61.4% (3,126)<sup>c</sup> | 58.6% (22,423)<sup>b,c</sup> |  |
| 3-7 | 16.0% (2,957)<sup>a,b</sup> | 18.7% (952)<sup>a</sup> | 18.7% (7,139)<sup>b</sup> |  |
| 8+ | 5.8% (1,077)<sup>b</sup> | 6.3% (320)<sup>c</sup> | 5.2% (2,002)<sup>b,c</sup> |  |
| Number of AUDIT-C assessments – mean (SD) | 9.2 (4.3)<sup>a,b</sup> | 7.8 (4.1)<sup>a,c</sup> | 8.9 (4.3)<sup>b,c</sup> | <0.0001 |
| AUD | 14.9% (2,757)<sup>a,b</sup> | 13.0% (662)<sup>a</sup> | 13.1% (5,005)<sup>b</sup> | <0.0001 |
| Cirrhosis<sup>**</sup> | 0.3% (53) | 0.3% (17) | 0.4% (152) | 0.1150 |
| Neuropathy<sup>**</sup> | 0.1% (13) | 0.0% (2) | 0.1% (31) | 0.5740 |
| Cardiomyopathy<sup>**</sup> | 0.0% (8) | 0.0% (0) | 0.0% (8) | 0.1589 |
| Gastritis<sup>**</sup> | 0.1% (20) | 0.1% (5) | 0.1% (44) | 0.9326 |
| Fatty liver disease<sup>**</sup> | 0.1% (24) | 0.3% (14) | 0.2% (74) | 0.0651 |
| Hepatitis<sup>**</sup> | 0.2% (33) | 0.2% (12) | 0.3% (103) | 0.1182 |
| Liver damage<sup>**</sup> | 0.1% (26) | 0.1% (5) | 0.2% (57) | 0.6636 |
| *Other Clinical ICD-9/10 Diagnoses* |  |  |  |  |
| Drug abuse/dependence | 13.2% (2,436)<sup>a,b</sup> | 9.7% (495)<sup>a,c</sup> | 11.2% (4,276)<sup>b,c</sup> | <0.0001 |
| AUD + drug abuse/dependence | 8.5% (1,572)<sup>a,b</sup> | 6.0% (304)<sup>a</sup> | 6.4% (2,452)<sup>b</sup> | <0.0001 |
| Mental disorder | 71.3% (13,154)<sup>a,b</sup> | 73.4% (3,739)<sup>a,c</sup> | 70.0% (26,773)<sup>b,c</sup> | <0.0001 |
| AUD + mental disorder | 14.1% (2,607)<sup>a,b</sup> | 12.5% (635)<sup>a</sup> | 12.4% (4,742)<sup>b</sup> | <0.0001 |

SD = standard deviation; AUDIT-C = Alcohol Use Disorders Identification Test – Consumption; AUD = alcohol use disorder.
\* Number of observations with missing age: Black men (n=5); Hispanic men (n=8); White men (n=42); Black women (n=1); Hispanic women (n=1); White women (n=3). \*\* Alcohol-specific diagnosis.
<sup>a</sup>Black vs Hispanic pairwise comparison significant at $P<0.05$ ; <sup>b</sup>Black vs White pairwise comparison significant at $P<0.05$ ; <sup>c</sup>Hispanic vs White pairwise comparison significant at $P<0.05$ .

#### Association between AUDIT-C and AUD by Race/Ethnicity

The correlation between AUDIT-C and AUD diagnosis was lowest among WMV (rho=0.36; *P*<0.0001), followed by HMV (rho=0.43; *P*<0.0001) and BMV (rho=0.47; *P*<0.0001). At every maximum AUDIT-C score, WMV were less likely than BMV to receive an AUD diagnosis (Figure 1a; all *P*<0.0001). The greatest difference was at AUDIT-C=4 (i.e., the positive screening cutoff for men), where WMV were approximately one-third as likely to have an AUD diagnosis as BMV. Although HMV had lower AUD frequency than BMV across all AUDIT-C scores, it was generally higher than among WMV, with the greatest difference also at AUDIT-C=4 (*P*<0.0001). The greatest difference between HMV and BMV was at AUDIT-C=7, where HMV were almost one-third less likely to have an AUD diagnosis than BMV (*P*<0.0001).

**Figure 1a.**
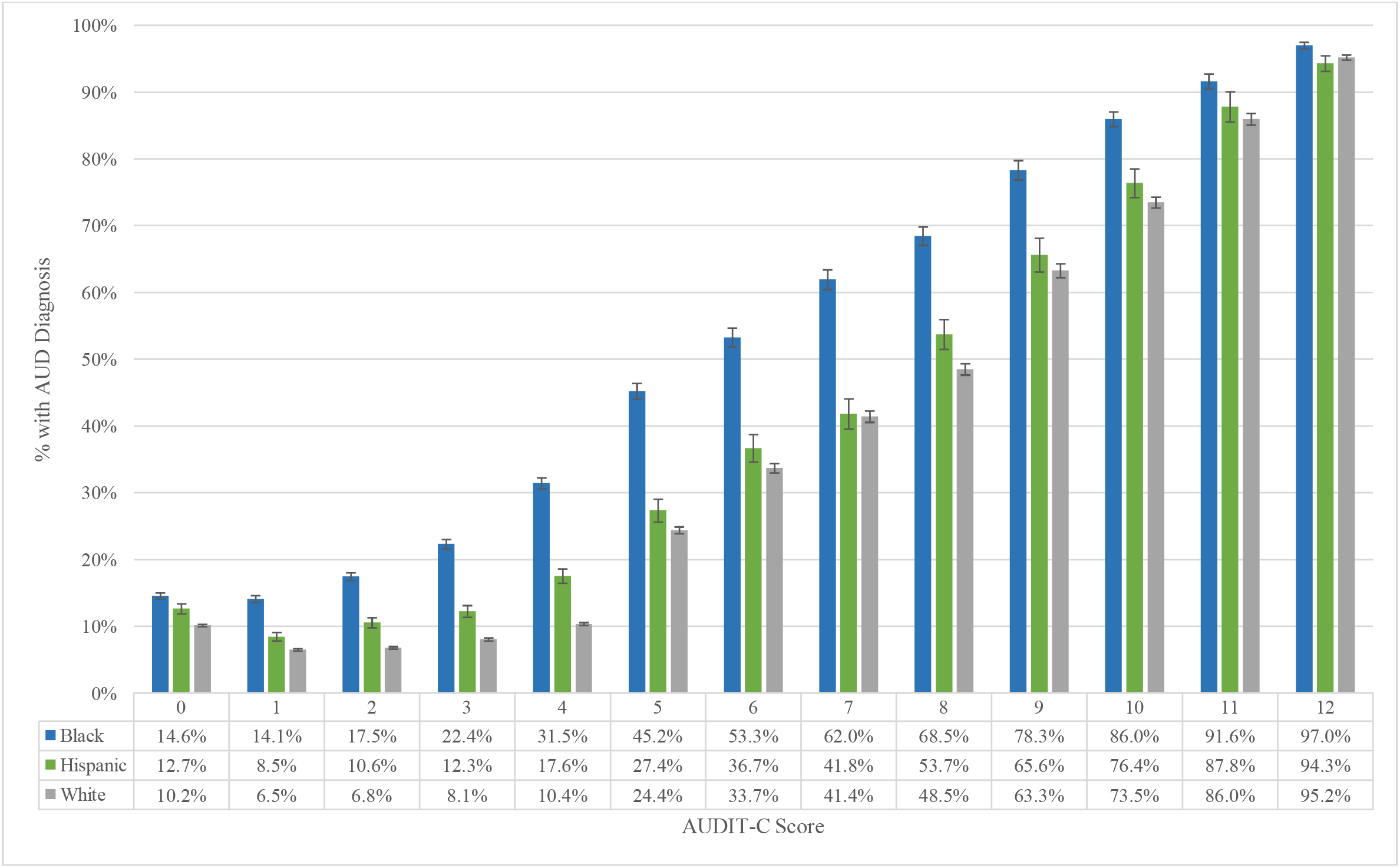
Percentage of men with an AUD diagnosis by maximum AUDIT-C score, stratified by race/ethnicity (n=638,205). AUD=alcohol use disorder; AUDIT-C=Alcohol Use Disorders Identification Test – Consumption.

**Figure 1b.**
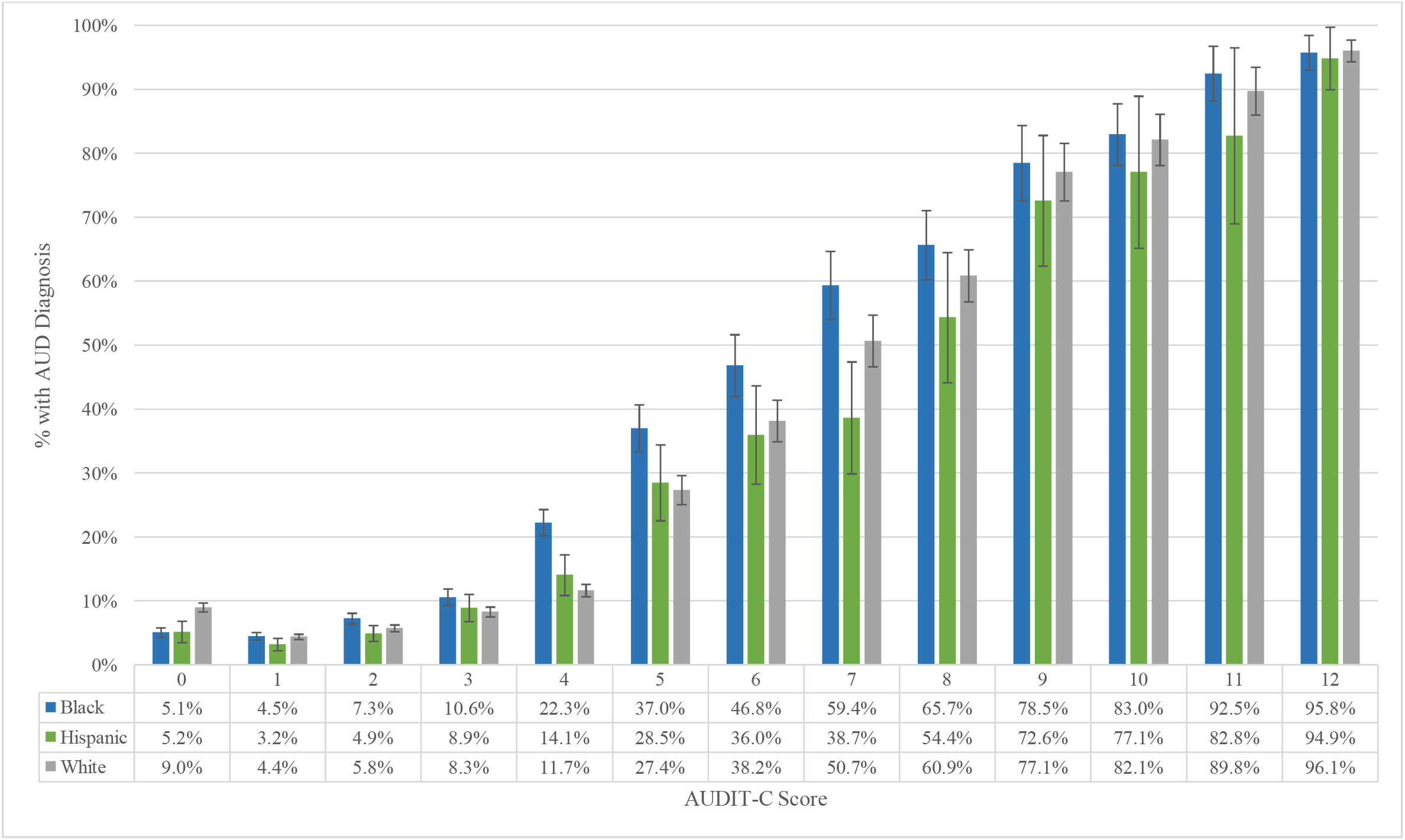
Percentage of women with an AUD diagnosis by maximum AUDIT-C score, stratified by race/ethnicity (n=61,808). AUD=alcohol use disorder; AUDIT-C=Alcohol Use Disorders Identification Test – Consumption.

BMV were significantly more likely than HMV and WMV to have an AUD diagnosis regardless of binge drinking frequency (all *P*<0.0001; Supplemental Figure 3a). Overall, HMV were more likely to have an AUD diagnosis than WMV regardless of binge drinking frequency.

#### Multivariable Analysis

Similar to findings from bivariate analyses, in multivariable analyses with adjustment for alcohol consumption and other potential confounders (Table 2), WMV were less likely to receive an AUD diagnosis than BMV and HMV. *Post hoc* analysis of the significant interaction between race/ethnicity and AUD diagnosis by alcohol consumption (Figure 2) revealed that BMV had 24%-111% greater odds of an AUD diagnosis than WMV at maximum AUDIT-C=1-10. At AUDIT-C=4, BMV had over twice the odds of an AUD diagnosis as WMV (*P*<0.0001). HMV were significantly more likely than WMV to have an AUD diagnosis at maximum AUDIT-C=2-4, with the highest odds at AUDIT-C=2 (*P*<0.0001).

**Table 2.** Factors associated with AUD diagnosis, overall and stratified by sex.

| *Variables* | Overall (n=699,953)* |  | Men (n=638,150)* |  | Women (n=61,803)* |  |
| --- | --- | --- | --- | --- | --- | --- |
|  | aOR (95% CI) | *P* | aOR (95% CI) | *P* | aOR (95% CI) | *P* |
| Race/Ethnicity x highest<br>AUDIT-C score** |  |  |  |  |  |  |
| Black (ref=White) | 1.93 (1.82, 2.04) | <0.0001 | 2.04 (1.92, 2.17) | <0.0001 | 1.22 (1.02, 1.45) | 0.0273 |
| Hispanic (ref=White) | 1.26 (1.15, 1.39) | <0.0001 | 1.28 (1.16, 1.41) | <0.0001 | 1.04 (0.77, 1.41) | 0.7983 |
| Women (ref=men) | 0.67 (0.65, 0.69) | <0.0001 | - | - | - | - |
| Age (10-yr increments) | 1.05 (1.05, 1.05) | <0.0001 | 1.05 (1.05, 1.06) | <0.0001 | 1.09 (1.08, 1.09) | <0.0001 |
| *Alcohol-Related Characteristics* |  |  |  |  |  |  |
| Cirrhosis*** | 15.02 (13.94, 16.17) | <0.0001 | 14.84 (13.77, 16.00) | <0.0001 | 19.41 (11.68, 32.24) | <0.0001 |
| Neuropathy*** | 14.35 (11.53, 17.87) | <0.0001 | 13.80 (11.08, 17.19) | <0.0001 | 176.80 (17.68,<br>>999.99) | <0.0001 |
| Cardiomyopathy*** | 15.91 (12.83, 19.72) | <0.0001 | 15.71 (12.66, 19.50) | <0.0001 | 32.55 (4.89, 216.94) | 0.0003 |
| Gastritis*** | 30.70 (19.92, 47.30) | <0.0001 | 31.11 (19.89, 48.66) | <0.0001 | 21.67 (4.07, 115.46) | 0.0003 |
| Fatty liver disease*** | 4.57 (4.06, 5.14) | <0.0001 | 4.59 (4.08, 5.18) | <0.0001 | 4.31 (2.31, 8.05) | <0.0001 |
| Hepatitis*** | 14.41 (11.47, 18.11) | <0.0001 | 15.11 (11.94, 19.12) | <0.0001 | 4.99 (1.97, 12.61) | 0.0007 |
| Liver damage*** | 8.38 (6.82, 10.29) | <0.0001 | 8.04 (6.52, 9.90) | <0.0001 | 22.01 (7.24, 66.97) | <0.0001 |
| *Other Clinical ICD-9/10 Diagnoses* |  |  |  |  |  |  |
| Drug use disorder | 12.82 (12.57, 13.08) | <0.0001 | 12.70 (12.44, 12.97) | <0.0001 | 13.11 (12.22, 14.07) | <0.0001 |
| Mental disorder | 3.19 (3.14, 3.25) | <0.0001 | 3.14 (3.08, 3.20) | <0.0001 | 5.06 (4.51, 5.67) | <0.0001 |
| C-statistic | 0.91 |  | 0.91 |  | 0.91 |  |

aOR=adjusted odds ratio; 95% CI =95% confidence interval; AUDIT-C=Alcohol Use Disorders Identification Test – Consumption;
AUD=alcohol use disorder. \* Number of observations with missing age: Black men (n=5); Hispanic men (n=8); White men (n=42);
Black women (n=1); Hispanic women (n=1); White women(n=3). \*\* Odds ratios displayed at mean highest AUDIT-C score (mean=3,
3, 2 for overall, men, and women, respectively). \*\*\* Alcohol-specific diagnosis.

**Figure 2.**
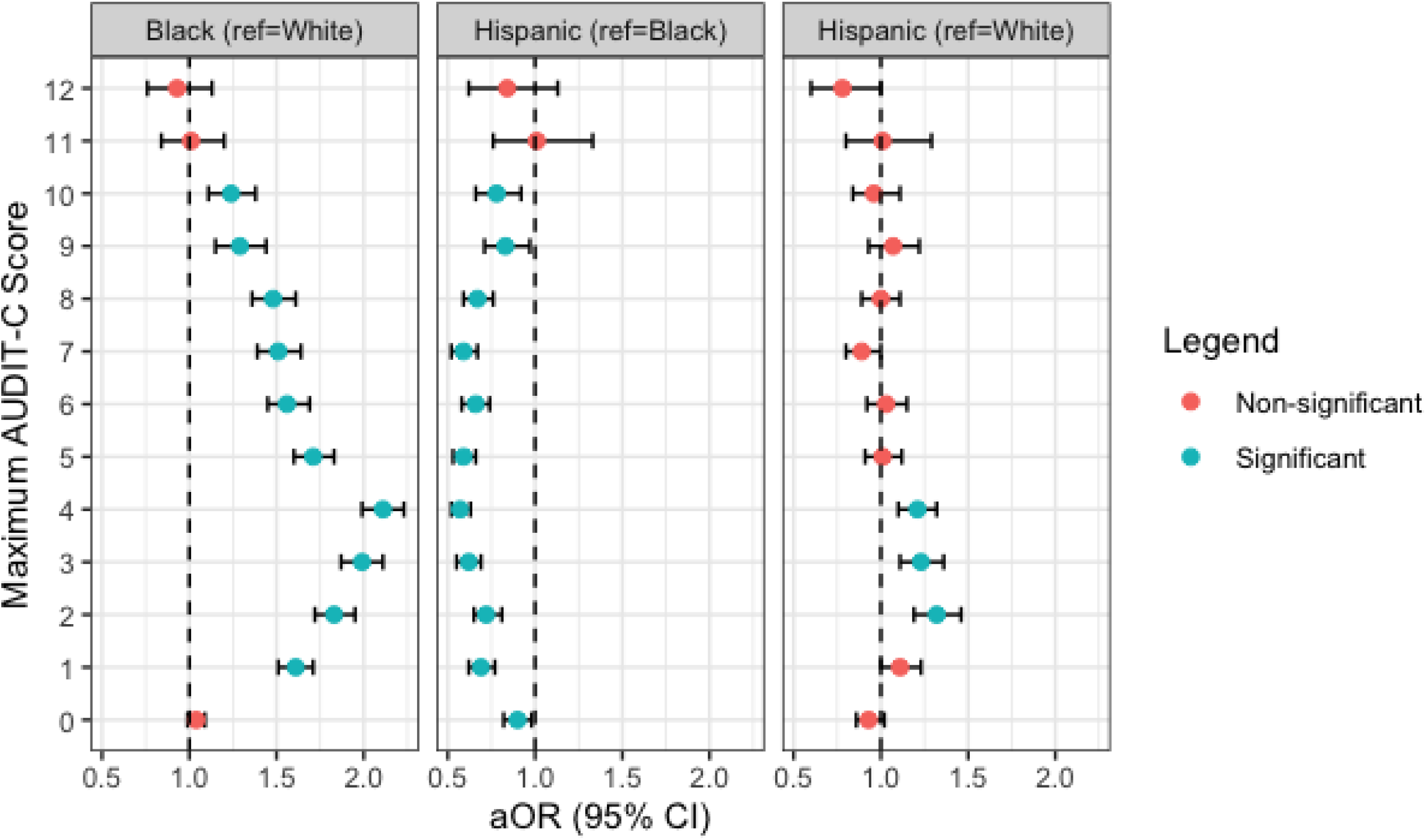
Forest plot of adjusted odds ratios for the association between race/ethnicity and AUD diagnosis among men, stratified by maximum AUDIT-C score (n=612,112). AUD=alcohol use disorder; AUDIT-C=Alcohol Use Disorders Identification Test – Consumption; aOR=adjusted odds ratio; 95% CI=95% confidence interval. Note: Models are adjusted for age at enrollment; drug abuse/dependence; mental disorder; alcohol-specific: cirrhosis, neuropathy, cardiomyopathy, gastritis, fatty liver disease, hepatitis, liver damage.

Among men, having a drug use disorder diagnosis was associated with nearly 13-fold increased odds of an AUD diagnosis (*P*<0.0001). Greater age, a mental disorder diagnosis, and alcohol-related medical disorders were also associated with significantly greater odds of an AUD diagnosis (*P*<0.0001).

### Women

#### Sample Characteristics

The racial/ethnic distribution among women was White (62%), Black (30%), and Hispanic (8%; Table 1). The modal maximal AUDIT-C score range was 1-2 for each group. Black female Veterans (BFV) had more AUDIT-C assessments on average (mean=9.2; SD=4.3) than Hispanic (HFV; mean=7.8; SD=4.1) and White (WFV; mean=8.9; SD=4.3) female Veterans. The frequency of an AUD diagnosis among BFV (15%) was significantly higher than WFV or HFV (both 13%; *P*<0.0001 and *P*=0.001, respectively). BFV had the highest frequency (13%) and HFV the lowest frequency (10%) of a drug use disorder. HFV were more likely to have a mental disorder diagnosis (73%) than BFV (71%) and WFV (70%). There were no significant racial/ethnic group differences in the prevalence of alcohol-related diagnoses.

#### Association between AUDIT-C and AUD by Race/Ethnicity

The correlation between AUDIT-C and AUD diagnosis was lower among WFV (rho=0.30; *P*<0.0001) than HFV (rho=0.34; *P*<0.0001) or BFV (rho=0.40; *P*<0.0001). BFV had the highest percentage of AUD diagnosis at nearly every level of alcohol consumption (Figure 1b) and were significantly more likely than WFV and HFV to receive an AUD diagnosis at AUDIT-C=2 and 4-7 (all *P*<0.05). At AUDIT-C=0, 1, and 7, WFV were more likely to have an AUD diagnosis than HFV. The maximal difference in the proportion of AUD diagnoses between BFV and WFV was at AUDIT-C=4 (22% vs. 12%; *P*<0.0001).

Racial/ethnic differences in the association between AUD frequency of binge drinking were less evident in women than men. BFV were more likely to receive an AUD diagnosis than HFV when reporting binge drinking weekly or less often (AUDIT-C question 3 scores=0-3; all *P*≤0.01; Supplemental Figure 3b).

#### Multivariable Analysis

The overall interaction between race/ethnicity and maximum AUDIT-C was significant (*P*<0.0001). BFV had a higher probability of an AUD diagnosis compared to HFV and WFV at moderate AUDIT-C scores and HFV had a lower probability of an AUD diagnosis compared to BFV and WFV at higher AUDIT-C scores (Supplemental Figure 4, Table 2). A drug use disorder diagnosis was associated with over 13-fold increase in the odds of an AUD diagnosis (*P*<0.0001). Greater age, a mental disorder diagnosis, and alcohol-related health conditions were associated with significantly greater odds of an AUD diagnosis (all *P*≤0.001).

### Sensitivity Analysis Using Age-Adjusted Mean AUDIT-C Score

Using age-adjusted mean rather than maximal AUDIT-C score did not affect the findings overall or by sex (Supplemental Table 3).

## Discussion

In this national cohort study of 700,000 veterans, we identified a differential frequency of AUD diagnosis by race/ethnicity. The greatest discrepancy was among Black men who, at all but the lowest and highest levels of alcohol consumption, had 24%-111% greater odds of an AUD diagnosis than White men. Hispanic men had 21%-32% greater odds of an AUD diagnosis than White men. The prevalence of disorders associated with persistent heavy drinking (e.g., alcoholic cirrhosis and hepatitis), whose diagnosis generally relies on objective measures (e.g., laboratory values, ultrasound findings), was similar across the three groups, indicating AUD diagnosis among Black and Hispanic Veterans was not due to greater alcohol consumption than White veterans. The difference in AUD diagnosis across the groups is also not attributable to the slightly higher percentages of positive AUDIT-C screens among Black and Hispanic Veterans than White Veterans. Further, the association between race/ethnicity and AUD diagnosis remained after adjustment for alcohol consumption, alcohol-related disorders, drug use disorder, and other potential confounders.

This study used EHR data on self-reported alcohol consumption and AUD diagnosis in a sample large enough to account for a variety of potential contributing factors. The AUD frequency across race/ethnicity and sex is similar to that previously observed in the Veteran population.^11^ Together, these findings highlight differential clinical assessment of AUD by race/ethnicity. It is unclear whether the difference is due to an overdiagnosis of Black Veterans, underdiagnosis of White Veterans, or a combination of the two, both of which are harmful, as overdiagnosis can be stigmatizing and underdiagnosis can delay treatment. The inequitable diagnostic process occurs in the context of Black Veterans being referred and receiving alcohol treatment at higher rates than White or Hispanic Veterans,^12,13^ though it is unclear whether this results in better outcomes or what the potential impact is of receiving an AUD diagnosis. Further, any potential benefit of disparate treatment rates should not overshadow the central issue that racial/ethnic inequity in assessment, particularly toward Black patients, appears to exist. Studies are needed to examine the mechanism by which Veterans receive an AUD diagnosis and factors that likely affect the observed inequity such as implicit bias and systemic racism.

The greatest disparity in AUD diagnosis after adjustment for potential confounders occurred at a maximum AUDIT-C score of 3 or 4, near the cutoff for a positive AUDIT-C screen (≥4 for men; ≥3 for women). This suggests that at scores near the threshold, providers are more likely to assign a diagnosis to Black or Hispanic Veterans than White Veterans.^7,24,25^ In a series of experiments that evaluated implicit stereotyping, physicians were more likely to associate negative stereotypes (e.g., drug abuse, HIV) with Black than White patients,^7,24^ suggesting that diagnostic disparities may be due to implicit bias. Some studies have suggested that diagnostic disparities could be due to the differential presentation of psychiatric symptoms across racial and ethnic groups.^11,26^ While this is possible due to the impact of culture on psychiatric symptom presentation,^27^ it suggests that diagnostic practice references the White experience. In this study, no data were available on why a diagnosis was given, although future studies could utilize diagnostic interviews to learn whether there are differences in endorsed criteria across race/ethnicity.

We found that the presence of a drug use disorder diagnosis was highly correlated with the diagnosis of AUD. These disorders commonly co-occur, both in the VA^28^ and the general population.^29,30^ However, in nationally representative epidemiologic studies, the prevalence of concurrent alcohol and drug *use* does not differ substantially by race/ethnicity (Black=9%, Hispanic=7%, White=8%),^30^ suggesting that alcohol and drug use disorder *diagnoses* should not differ substantially by race/ethnicity. However, in this sample, Black men were over three times and Hispanic men were nearly twice as likely to have at least one comorbid drug diagnosis as White men. One explanation for these findings could be that once a patient receives an AUD diagnosis, providers may be more likely to query the patient about other substance use. Findings may also be due to implicit bias toward Black and Hispanic Veterans resulting in additional screening for additional substance use in these racial/ethnic populations.^7,31,32^ More research is needed to understand the source of these differences. Further, to aid equitable diagnosis and treatment of multiple substance use (beyond alcohol and tobacco), standardized screening and assessment are recommended.

Women were less likely than men to have an AUD diagnosis, consistent with population estimates^33^ and findings among Veterans.^34^ Although women consume less alcohol than men, the sex/gender difference has been decreasing in recent years.^35^ In studies of unhealthy alcohol use, women experience greater alcohol use related stigma than men,^36,37^ which could impact how providers respond to^38,39^ and document^40^ use among women. More research is needed to understand sex/gender differences in substance use reporting and its documentation in the EHR.

As seen among men, at nearly every level of alcohol consumption, Black women were more likely than Hispanic and White women to receive an AUD diagnosis, despite the groups’ similar distributions of alcohol consumption and prevalence of alcohol-related disorders. There were few differences in the relation between AUD frequency and alcohol consumption between Hispanic and White women. Where such differences occurred, White women had a greater AUD frequency than Hispanic women, consistent with estimates from the 2019 National Survey of Drug Use and Health where alcohol use prevalence is higher among non-Hispanic White women than Hispanic women.^33^

This study has several limitations. Despite obvious differences in the frequency of an AUD diagnosis by racial/ethnic group, the basis for the discrepancies cannot be ascertained using EHR data. Second, measures of alcohol consumption relied on self-report, which may not be valid. In future studies alcohol biomarkers could serve as objective measures of alcohol use. Lastly, findings from this study of US veterans enrolled in a genetic cohort study may not generalize to other populations, even the general Veteran population.

Our study also has a number of notable strengths. The availability of annual assessments of alcohol consumption and the availability of an informative EHR enabled the ability to analyze relationships between measures of alcohol consumption and an AUD diagnosis, as well as multiple clinical factors that could influence that association. Second, the large and diverse sample made it possible to examine factors that could affect the likelihood of an AUD diagnosis.

We identified a large difference in AUD diagnosis by race/ethnicity, which was discrepant with self-reported alcohol consumption. As other factors do not appear to explain this discrepancy, the findings suggest that there is bias in the diagnosis of AUD by race/ethnicity. These findings should encourage the VA and other healthcare systems to examine the causes of any observed differences, and to implement changes, such as the use of structured diagnostic methods, to address a likely contributor to racial differences in AUD diagnosis (i.e., bias).

## Data Availability

Data described in the manuscript will not be made available to other researchers for purposes of reproducing the results or replicating the procedure, in order to comply with current VA privacy regulations pursuant to the US Department of Veterans Administration policies on compliance with the confidentiality of US Veterans' data.

## Acknowledgements

We thank all US Veterans for their service and particularly those Veterans who participated in the Million Veteran Program. This research is based on data from the Million Veteran Program, Office of Research and Development, Veterans Health Administration, and was supported by award #I01 BX003341 and the Veterans Integrated Service Network 4 Mental Illness Research, Education and Clinical Center. This publication does not represent the views of the Department of Veteran Affairs or the United States Government. This research was supported in part by the Intramural Research Program of the NIH, NIDA.

