## Supplemental Appendix for "Racial Inequalities in Alcohol Use Disorder Diagnosis in a Sample of 700,000 Veterans"

### Supplementary Appendix

*Study Sample*

The study sample was drawn from the Million Veteran Program (MVP), an ongoing longitudinal cohort of US veterans (described in detail in[1]). Veterans who receive care in the VA and consented to participate in the MVP were asked to complete two self-report surveys and provide access to their electronic health records (EHR). MVP enrollment began in early 2011 and as of December 2020 more than 800,000 veterans were enrolled. Of this number, we had access to data for 790,091, of whom 739,412 had AUDIT-C, race/ethnicity, and sex data available in their EHR (see Supplemental Figure 1 for a flow diagram of study inclusion). Of those, 700,013 were included in the primary analysis as described in the race/ethnicity section below.

*Measures*

*Race/ethnicity.* Race/ethnicity was self-reported in both an MVP questionnaire and the VA EHR; when race/ethnicity was missing from the MVP survey, data from the EHR was used.[2] We focus here on three groups: non-Hispanic White (White), non-Hispanic Black (Black), and Hispanic. Participants who self-identified as Hispanic were classified as Hispanic irrespective of racial designation consistent with the U.S. Census Bureau.[3] Other racial/ethnic groups (including multi-racial), information on which is presented in supplementary analyses (see Supplemental

---

[1] Gaziano JM, Concato J, Brophy M, et al. Million Veteran Program: A mega-biobank to study genetic influences on health and disease. *J Clin Epidemiol.* 2016;70:214-223. Doi:10.1016/j.clinepi.2015.09.016

[2] Harrington KM, Nguyen XMT, Song RJ, et al. Gender differences in demographic and health characteristics of the Million Veteran Program cohort. *Women's Heal Issues.* 2019;29(Suppl 1):S56-S66. Doi:10.1016/j.whi.2019.04.012

[3] Hispanic Origin. https://www.census.gov/topics/population/hispanic-origin.html. Accessed April 9, 2021.

Tables 1a & b, Figures 2a & b), were excluded from the primary analyses because they were much smaller than the three primary groups.

*Self-reported alcohol consumption.* The AUDIT-C is a valid, reliable screening instrument that is routinely used to identify individuals who engage in hazardous or harmful drinking.[4,5] It consists of the first 3 items of the 10-item AUDIT, which measures past-year alcohol consumption, as follows:[6] (i) How often do you have a drink containing alcohol? (Drinking Frequency–response options: never, monthly or less, 2 to 4 times a month, 2 to 3 times a week, 4 or more times a week); (ii) How many standard drinks containing alcohol do you have on a typical day? (Drinking Quantity–response options: 1 or 2, 3 or 4, 5 or 6, 7 to 9, 10 or more); and (iii) How often do you have 6 or more drinks on 1 occasion? (Binge Drinking Frequency–response options: never, less than monthly, monthly, weekly, daily or almost daily). The responses to each question are scored 0–4 and the points from individual questions are summed for a total AUDIT-C score of 0–12, with higher scores indicating greater risk for hazardous drinking. AUDIT-C scores of ≥3 for women and ≥4 for men reflect hazardous drinking[7] and

---

[4] Bush K, Kivlahan DR, McDonnell MB, Fihn SD, Bradley KA. The AUDIT alcohol consumption questions (AUDIT-C): An effective brief screening test for problem drinking. *Arch Intern Med.* 1998;158(16):1789. Doi:10.1001/archinte.158.16.1789

[5] Bradley KA, Debenedetti AF, Volk RJ, Williams EC, Frank D, Kivlahan DR. AUDIT-C as a brief screen for alcohol misuse in primary care. *Alcohol Clin Exp Res.* 2007;31(7):1208-1217. Doi:10.1111/j.1530-0277.2007.00403.x

[6] Saunders JB, Aasland OG, Babor TF, De La Fuente JR, Grant M. Development of the Alcohol Use Disorders Identification Test (AUDIT): WHO collaborative project on early detection of persons with harmful alcohol consumption-II. *Addiction.* 1993;88(6):791-804. Doi:10.1111/j.1360-0443.1993.tb02093.x

[7] See note 5 above

indicate further assessment is needed. When used to identify individuals with current DSM-IV[8] alcohol abuse or dependence, the AUDIT-C at these cutoffs had a sensitivity/ specificity of 0.79/0.56 among men[9] and 0.80/0.87 among women.[10] By race/ethnicity, when used to identify individuals with alcohol use disorders or risky drinking, the AUDIT-C at those cutoffs had a sensitivity/specificity of 0.67/0.92 among African American women, 0.85/0.88 among Hispanic women, 0.70/0.91 among White women, 0.76/0.93 among African American men, 0.85/0.84 among Hispanic men, and 0.95/0.89 among White men.[11]

To capture an individual's maximal reported alcohol consumption, we extracted the highest AUDIT-C recorded in their VA EHR (AUDIT-C dates ranged from December 9, 1999 to September 30, 2019). In sensitivity analyses, we used age-adjusted mean AUDIT-C, with age 50 as the reference point, and up-weighted AUDIT-C scores for individuals older than 50 and down-weighted scores for those younger than 50. Each AUDIT-C score was multiplied by the

---

[8] American Psychiatric Association. *Diagnostic and Statistical Manual of Mental Disorders.* 4th ed. Washington, DC: American Psychiatric Association; 1994. https://search.library.wisc.edu/catalog/999733358502121

[9] See note 4 above

[10] Bradley KA, Bush KR, Epler AJ, et al. Two brief alcohol-screening tests from the Alcohol Use Disorders Identification Test (AUDIT): Validation in a female Veterans Affairs patient population. *Arch Intern Med.* 2003;163(7):821-829. Doi:10.1001/archinte.163.7.821

[11] Frank D, DeBenedetti AF, Volk RJ, Williams EC, Kivlahan DR, Bradley KA. Effectiveness of the AUDIT-C as a screening test for alcohol misuse in three race/ethnic groups. *J Gen Intern Med.* 2008;23(6):781-787. Doi:10.1007/s11606-008-0594-0.

weight corresponding to age at the time of the AUDIT-C assessment and weighted AUDIT-C scores were summed and divided by the weights used for each individual.[12,13]

Binge drinking, the frequency of which is assessed by AUDIT-C item 3, is reportedly particularly harmful and indicative of greater vulnerability to AUD.[14] To evaluate whether, in addition to total AUDIT-C score, the relationship between race/ethnicity and AUD diagnosis is impacted by binge drinking frequency, we also examined the association between the maximum score on AUDIT-C item 3 and AUD diagnosis by race/ethnicity.

*Demographics and clinical diagnoses.* Sex (male or female) and age at enrollment (calculated from month and year of birth and date of MVP enrollment) were based on the MVP questionnaires and linked EHR data; as described above when sex or age was missing from the MVP survey, data from the EHR was used.[15] Clinical diagnoses required the presence of one inpatient or two outpatient ICD-9/10 diagnostic codes in the VA EHR and included AUD; alcohol-related disorders (cirrhosis, neuropathy, cardiomyopathy, gastritis, fatty liver disease, hepatitis, and liver damage); drug use disorder (abuse/dependence on opioids, cannabis, barbiturates, cocaine, amphetamines, and other stimulants, sedatives, and psychoactive

---

[12] Justice AC, Smith RV, Tate JP, et al. AUDIT-C and ICD codes as phenotypes for harmful alcohol use: Association with *ADH1B* polymorphisms in two US populations. *Addiction*. 2018;113(12):2214-2224. Doi:10.1111/add.14374

[13] Vickers Smith R, Kranzler HR, Justice AC, Tate JP, Million Veteran Program. Longitudinal drinking patterns and their clinical correlates in Million Veteran Program participants. *Alcohol Clin Exp Res.* 2019;43(3):465-472. Doi:10.1111/acer.13951

[14] Gowin JL, Sloan ME, Stangl BL, Vatsalya V, Ramchandani VA. Vulnerability for alcohol use disorder and rate of alcohol consumption. *Am J Psychiatry.* 2017;174(11):1094-1101. doi:10.1176/appi.ajp.2017.16101180

[15] See note 2 above

substances); and mental disorder (schizophrenia, schizoaffective disorders, bipolar disorder, PTSD, and other anxiety disorders) (see Supplemental Table 2 for the specific ICD codes).
