## Supplemental Tables and Figures for "Racial Inequalities in Alcohol Use Disorder Diagnosis in a Sample of 700,000 Veterans"

Supplemental Figure 1. Flow chart of study inclusion.

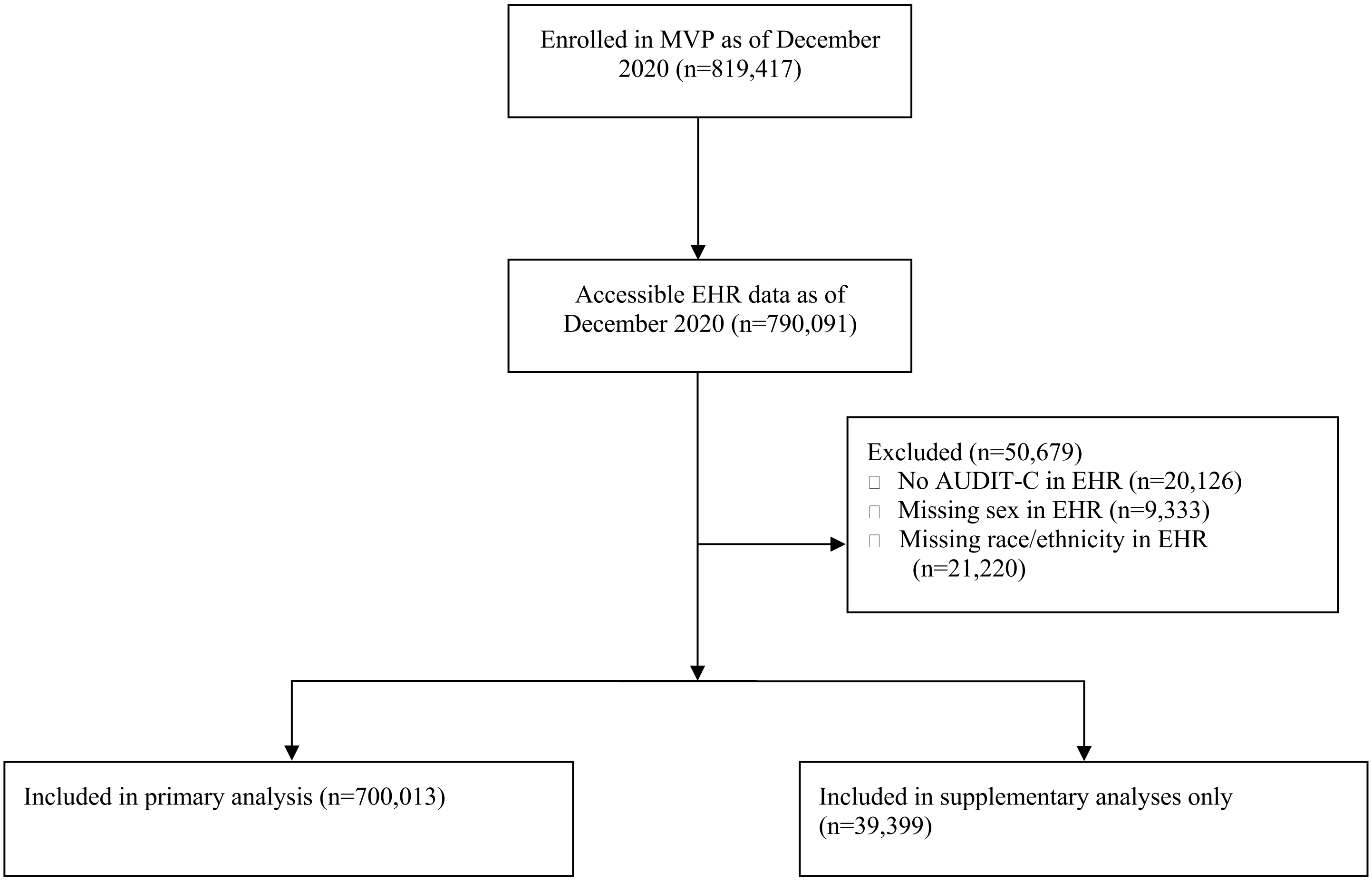


MVP = Million Veteran Program; EHR = electronic health record; AUDIT-C = Alcohol Use Disorders Identification Test – Consumption.

Supplemental Table 1a. Demographic and clinical characteristics of men excluded in the primary analysis by race/ethnicity (n=34,056).

| | American Indian/ Alaska Native 13.7% (n=4,677) | Asian 21.0% (n=7,135) | Native Hawaiian/ Pacific Islander 9.1% (n=3,102) | Other/Multiple Races 56.2% (n=19,142) |
|---|---|---|---|---|
| Age (y) – mean (SD)* | 60.1 (12.7) | 53.8 (17.0) | 57.3 (14.7) | 61.4 (12.5) |
| *Alcohol-Related Characteristics* | | | | |
| Highest AUDIT-C | | | | |
| 0 | 26.2% (1,223) | 20.7% (1,477) | 22.2% (689) | 22.5% (4,305) |
| 1-3 | 36.8% (1,721) | 49.5% (3,534) | 40.3% (1,250) | 43.8% (8,381) |
| 4-7 | 22.5% (1,050) | 21.8% (1,558) | 23.5% (729) | 23.2% (4,446) |
| 8+ | 14.6% (683) | 7.9% (566) | 14.0% (434) | 10.5% (2,010) |
| Number of AUDIT-C assessments – mean (SD) | 8.9 (4.4) | 7.1 (3.7) | 8.4 (4.0) | 9.1 (4.3) |
| AUD | 28.6% (1,336) | 11.7% (835) | 21.7% (672) | 20.9% (3,992) |
| Cirrhosis** | 2.5% (117) | 0.3% (19) | 1.6% (49) | 1.5% (283) |
| Neuropathy** | 0.2% (10) | 0.0% (0) | 0.1% (4) | 0.3% (56) |
| Cardiomyopathy** | 0.2% (10) | <0.1% (1) | 0.1% (2) | 0.2% (31) |
| Gastritis** | 0.3% (16) | 0.1% (5) | 0.1% (3) | 0.2% (43) |
| Fatty liver disease** | 0.6% (30) | 0.3% (19) | 0.8% (25) | 0.5% (92) |
| Hepatitis** | 0.9% (42) | 0.1% (8) | 0.6% (17) | 0.6% (113) |
| Liver damage** | 0.5% (24) | 0.1% (9) | 0.2% (7) | 0.3% (62) |
| *Other Clinical ICD-9/10 Diagnoses* | | | | |
| Drug abuse/dependence | 19.2% (897) | 6.5% (465) | 14.5% (451) | 15.4% (2,948) |
| AUD + drug abuse/dependence | 14.8% (694) | 4.2% (300) | 10.2% (317) | 10.8% (2,060) |
| Mental disorder | 62.0% (2,900) | 47.1% (3,359) | 60.0% (1,860) | 56.8% (10,878) |
| AUD + mental disorder | 23.7% (1,106) | 9.9% (703) | 18.0% (557) | 17.1% (3,279) |

SD = standard deviation; AUDIT-C = Alcohol Use Disorders Identification Test – Consumption; AUD = alcohol use disorder.
*Number of observations with missing age: Asian (n=4), Other/Multiple Races (n=2). **Alcohol-specific diagnosis.

Supplemental Table 1b. Demographic and clinical characteristics of women excluded in the primary analysis by race/ethnicity (n=5,343).

| | American Indian/ Alaska Native 13.1% (n=702) | Asian 17.5% (n=934) | Native Hawaiian/ Pacific Islander 7.4% (n=394) | Other/Multiple Races 62.0% (n=3,313) |
|---|---|---|---|---|
| Age (y) – mean (SD)* | 50.4 (12.8) | 43.1 (12.8) | 47.1 (12.7) | 51.4 (12.6) |
| *Alcohol-Related Characteristics* | | | | |
| Highest AUDIT-C | | | | |
| 0 | 21.7% (152) | 20.6% (192) | 20.6% (81) | 18.9% (626) |
| 1-3 | 54.3% (381) | 61.7% (576) | 59.4% (234) | 61.1% (2,024) |
| 4-7 | 18.4% (129) | 14.5% (135) | 15.0% (59) | 15.2% (504) |
| 8+ | 5.7% (40) | 3.3% (31) | 5.1% (20) | 4.8% (159) |
| Number of AUDIT-C assessments – mean (SD) | 8.7 (5.2) | 7.1 (3.6) | 8.4 (4.3) | 9.2 (4.4) |
| AUD | 17.1% (120) | 7.9% (74) | 13.2% (52) | 12.2% (404) |
| Cirrhosis** | 0.4% (3) | 0.0% (0) | 0.5% (2) | 0.4% (14) |
| Neuropathy** | 0.1% (1) | 0.0% (0) | 0.3% (1) | <0.1% (1) |
| Cardiomyopathy** | 0.1% (1) | 0.0% (0) | 0.0% (0) | 0.0% (0) |
| Gastritis** | 0.1% (1) | 0.0% (0) | 0.5% (2) | 0.1% (3) |
| Fatty liver disease** | 0.4% (3) | 0.0% (0) | 0.3% (1) | 0.2% (6) |
| Hepatitis** | 0.3% (2) | 0.0% (0) | 0.3% (1) | 0.2% (8) |
| Liver damage** | 0.6% (4) | 0.0% (0) | 0.3% (1) | 0.1% (2) |
| *Other Clinical ICD-9/10 Diagnoses* | | | | |
| Drug abuse/dependence | 15.5% (109) | 6.2% (58) | 11.4% (45) | 12.0% (399) |
| AUD + drug abuse/dependence | 9.5% (67) | 3.8% (35) | 5.6% (22) | 6.3% (210) |
| Mental disorder | 75.6% (531) | 59.9% (559) | 73.6% (290) | 75.0% (2,485) |
| AUD + mental disorder | 16.4% (115) | 7.6% (71) | 12.4% (49) | 11.6% (383) |

SD = standard deviation; AUDIT-C = Alcohol Use Disorders Identification Test – Consumption; AUD = alcohol use disorder.
*Number of observations with missing age: Other/Multiple Races (n=1). **Alcohol-specific diagnosis.

Supplemental Figure 2a. Percentage of men with an AUD diagnosis by maximum AUDIT-C score, by race/ethnicity (n=672,261).

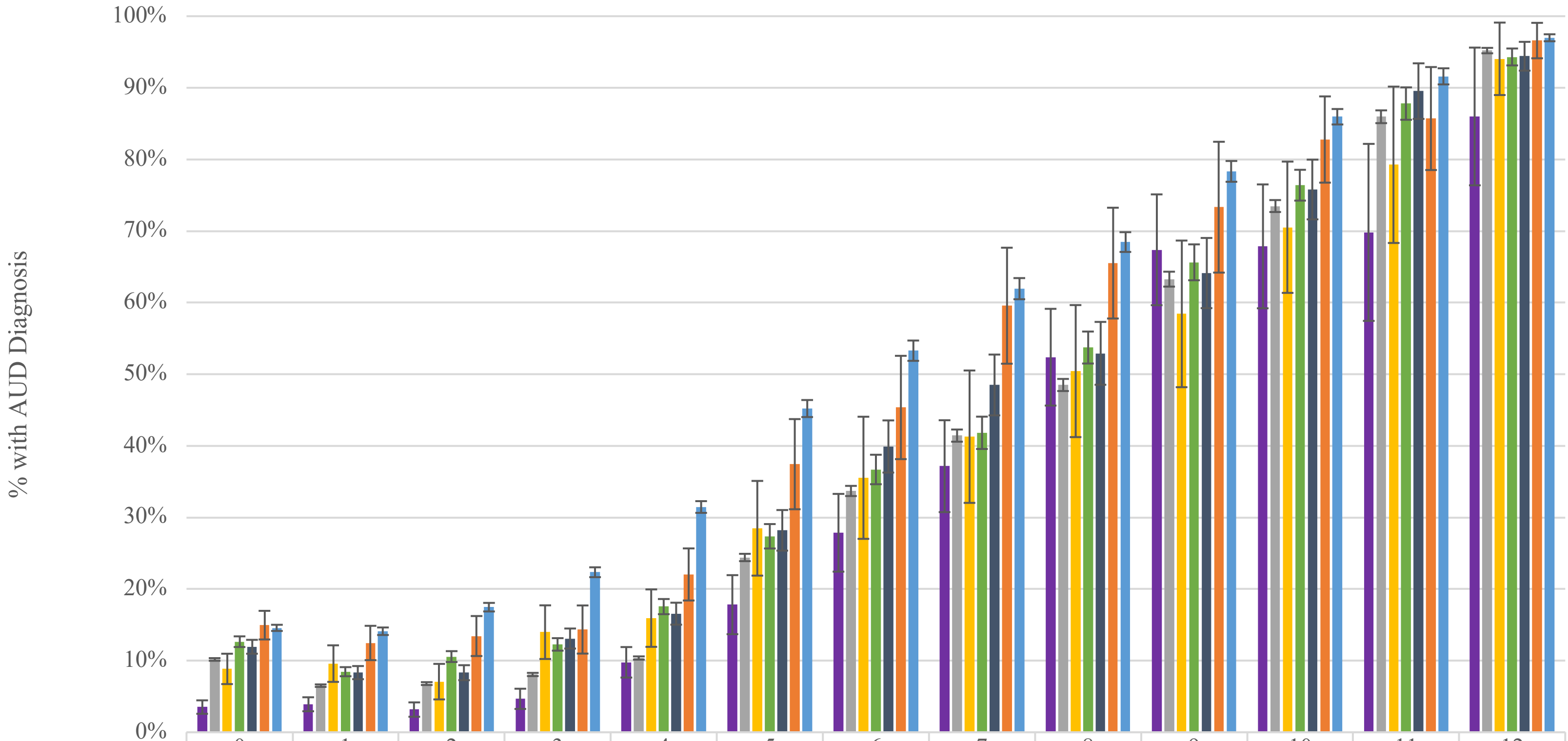


| | 0 | 1 | 2 | 3 | 4 | 5 | 6 | 7 | 8 | 9 | 10 | 11 | 12 |
|---|---|---|---|---|---|---|---|---|---|---|---|---|---|
| Asian | 3.5% | 3.9% | 3.2% | 4.7% | 9.8% | 17.8% | 27.9% | 37.2% | 52.4% | 67.4% | 67.9% | 69.8% | 86.0% |
| White | 10.2% | 6.5% | 6.8% | 8.1% | 10.4% | 24.4% | 33.7% | 41.4% | 48.5% | 63.3% | 73.5% | 86.0% | 95.2% |
| NH/PI | 8.9% | 9.6% | 7.1% | 14.0% | 15.9% | 28.5% | 35.5% | 41.3% | 50.4% | 58.4% | 70.5% | 79.3% | 94.1% |
| Hispanic | 12.7% | 8.5% | 10.6% | 12.3% | 17.6% | 27.4% | 36.7% | 41.8% | 53.7% | 65.6% | 76.4% | 87.8% | 94.3% |
| Other/Multiple | 11.9% | 8.3% | 8.3% | 13.1% | 16.6% | 28.2% | 39.9% | 48.5% | 52.9% | 64.1% | 75.8% | 89.5% | 94.4% |
| AI/AN | 15.0% | 12.5% | 13.4% | 14.4% | 22.0% | 37.4% | 45.4% | 59.6% | 65.5% | 73.3% | 82.8% | 85.7% | 96.6% |
| Black | 14.6% | 14.1% | 17.5% | 22.4% | 31.5% | 45.2% | 53.3% | 62.0% | 68.5% | 78.3% | 86.0% | 91.6% | 97.0% |

AUDIT-C Score

AUD=alcohol use disorder; AUDIT-C=Alcohol Use Disorders Identification Test – Consumption; NH/PI=Native Hawaiian/Pacific Islander; AI/AN=American Indian/Alaska Native.

Supplemental Figure 2b. Percentage of women with an AUD diagnosis by maximum AUDIT-C score, by race/ethnicity (n=67,151).

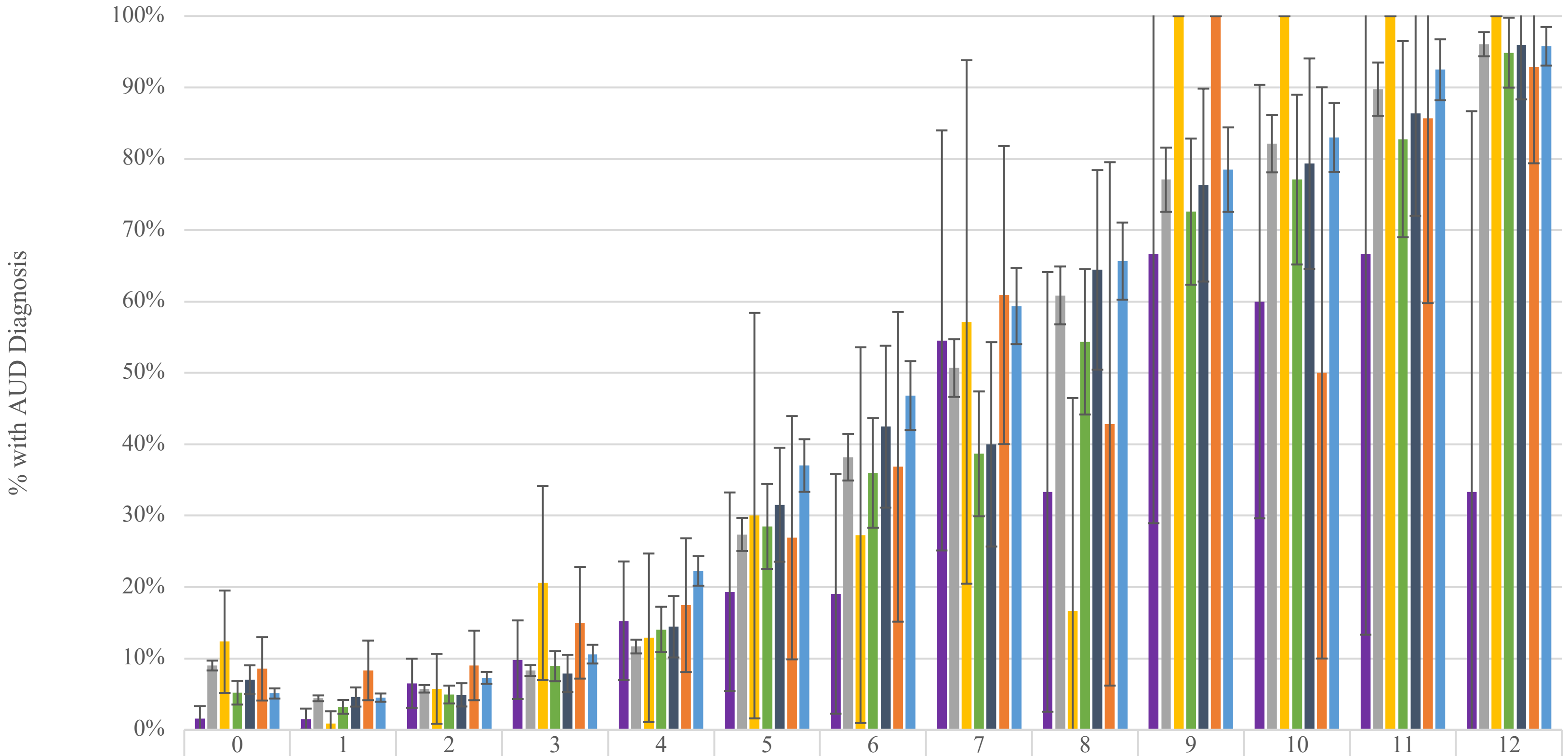


| | 0 | 1 | 2 | 3 | 4 | 5 | 6 | 7 | 8 | 9 | 10 | 11 | 12 |
|---|---|---|---|---|---|---|---|---|---|---|---|---|---|
| Asian | 1.6% | 1.5% | 6.5% | 9.8% | 15.3% | 19.4% | 19.1% | 54.6% | 33.3% | 66.7% | 60.0% | 66.7% | 33.3% |
| White | 9.0% | 4.4% | 5.8% | 8.3% | 11.7% | 27.4% | 38.2% | 50.7% | 60.9% | 77.1% | 82.1% | 89.8% | 96.1% |
| NH/PI | 12.4% | 0.9% | 5.8% | 20.6% | 12.9% | 30.0% | 27.3% | 57.1% | 16.7% | 100.0% | 100.0% | 100.0% | 100.0% |
| Hispanic | 5.2% | 3.2% | 4.9% | 8.9% | 14.1% | 28.5% | 36.0% | 38.7% | 54.4% | 72.6% | 77.1% | 82.8% | 94.9% |
| Other/Multiple | 7.0% | 4.6% | 4.9% | 7.9% | 14.5% | 31.5% | 42.5% | 40.0% | 64.4% | 76.3% | 79.3% | 86.4% | 96.0% |
| AI/AN | 8.6% | 8.3% | 9.0% | 15.0% | 17.5% | 26.9% | 36.8% | 60.9% | 42.9% | 100.0% | 50.0% | 85.7% | 92.9% |
| Black | 5.1% | 4.5% | 7.3% | 10.6% | 22.3% | 37.0% | 46.8% | 59.4% | 65.7% | 78.5% | 83.0% | 92.5% | 95.8% |

AUDIT-C Score

AUD=alcohol use disorder; AUDIT-C=Alcohol Use Disorders Identification Test – Consumption; NH/PI=Native Hawaiian/Pacific Islander; AI/AN=American Indian/Alaska Native.

Supplemental Table 2. International Classification of Disease (ICD) 9 and 10 Diagnostic Codes.

| Alcohol Use Disorder | | | |
|---|---|---|---|
| ICD-9 Codes | | ICD-10 Codes | |
| 305.00 | Nondependent alcohol abuse, unspecified | F10.10 | Alcohol abuse, uncomplicated |
| 305.01 | Nondependent alcohol abuse, continuous | F10.120 | Alcohol abuse with intoxication, uncomplicated |
| 305.02 | Nondependent alcohol abuse, episodic | F10.121 | Alcohol abuse with intoxication, delirium |
| 305.03 | Nondependent alcohol abuse, in remission | F10.129 | Alcohol abuse with intoxication, unspecified |
| 303.00 | Acute alcoholic intoxication, unspecified | F10.14 | Alcohol abuse with alcohol-induced mood disorder |
| 303.01 | Acute alcoholic intoxication, continuous | F10.150 | Alcohol abuse with alcohol-induced psychotic disorder with delusions |
| 303.02 | Acute alcoholic intoxication, episodic | F10.151 | Alcohol abuse with alcohol-induced psychotic disorder with hallucinations |
| 303.03 | Acute alcoholic intoxication, in remission | F10.159 | Alcohol abuse with alcohol-induced psychotic disorder, unspecified |
| 303.90 | Other and unspecified alcohol dependence, unspecified | F10.180 | Alcohol abuse with alcohol-induced anxiety disorder |
| 303.91 | Other and unspecified alcohol dependence, continuous | F10.181 | Alcohol abuse with alcohol-induced sexual dysfunction |
| 303.92 | Other and unspecified alcohol dependence, episodic | F10.182 | Alcohol abuse with alcohol-induced sleep disorder |
| 303.93 | Other and unspecified alcohol dependence, in remission | F10.188 | Alcohol abuse with other alcohol-induced disorder |
| | | F10.19 | Alcohol abuse with unspecified alcohol-induced disorder |
| | | F10.20 | Alcohol dependence, uncomplicated |
| | | F10.21 | Alcohol dependence, in remission |
| | | F10.220 | Alcohol dependence with intoxication, uncomplicated |
| | | F10.221 | Alcohol dependence with intoxication delirium |
| | | F10.229 | Alcohol dependence with intoxication, unspecified |
| | | F10.230 | Alcohol dependence with withdrawal, uncomplicated |
| | | F10.231 | Alcohol dependence with withdrawal delirium |
| | | F10.232 | Alcohol dependence with withdrawal with perceptual disturbance |

| | | | |
|---|---|---|---|
| [redacted] | | F10.239 | Alcohol dependence with withdrawal, unspecified |
| | | F10.24 | Alcohol dependence with alcohol-induced mood disorder |
| | | F10.250 | Alcohol dependence with alcohol-induced psychotic disorder with delusions |
| | | F10.251 | Alcohol dependence with alcohol-induced psychotic disorder with hallucinations |
| [redacted] | | F10.259 | Alcohol dependence with alcohol-induced psychotic disorder, unspecified |
| | | F10.26 | Alcohol dependence with alcohol-induced persisting amnestic disorder |
| | | F10.27 | Alcohol dependence with alcohol-induced persisting dementia |
| | | F10.280 | Alcohol dependence with alcohol-induced anxiety disorder |
| | | F10.281 | Alcohol dependence with alcohol-induced sexual dysfunction |
| | | F10.282 | Alcohol dependence with alcohol-induced sleep disorder |
| | | F10.288 | Alcohol dependence with other alcohol-induced disorder |
| | | F10.29 | Alcohol dependence with unspecified alcohol-induced disorder |
| **Alcohol-Related Disorders** | | | |
| **ICD-9 Codes** | | **ICD-10 Codes** | |
| 571.2 | Alcoholic cirrhosis of liver | K70.3 | Alcoholic cirrhosis of liver |
| [redacted] | | K70.30 | Alcoholic cirrhosis of liver without ascites |
| | | K70.31 | Alcoholic cirrhosis of liver with ascites |
| 357.5 | Alcoholic polyneuropathy | G62.1 | Alcoholic polyneuropathy |
| 425.5 | Alcoholic cardiomyopathy | I42.6 | Alcoholic cardiomyopathy |
| 535.3 | Alcoholic gastritis | K29.2 | Alcoholic gastritis |
| 535.30 | Alcoholic gastritis without mention of hemorrhage | K29.20 | Alcoholic gastritis without bleeding |
| 535.31 | Alcoholic gastritis with hemorrhage | K29.21 | Alcoholic gastritis with bleeding |
| 571.0 | Alcoholic fatty liver | K70.0 | Alcoholic fatty liver |

| | | | |
|---|---|---|---|
| 571.1 | Acute alcoholic hepatitis | K70.1 | Alcoholic hepatitis |
| [redacted] | | K70.10 | Alcoholic hepatitis without ascites |
| | | K70.11 | Alcoholic hepatitis with ascites |
| 571.3 | Alcoholic liver damage, unspecified | K70.9 | Alcoholic liver disease, unspecified |
| **Drug Abuse or Dependence** | | | |
| **ICD-9 Codes** | | **ICD-10 Codes** | |
| 304 | Drug dependence | F11.1 | Opioid abuse |
| 304.0 | Opioid type dependence | F11.10 | Opioid abuse, uncomplicated |
| 304.00 | Opioid type dependence, unspecified | F11.11 | Opioid abuse, in remission |
| 304.01 | Opioid type dependence, continuous | F11.12 | Opioid abuse with intoxication |
| 304.02 | Opioid type dependence, episodic | F11.120 | Opioid abuse with intoxication, uncomplicated |
| 304.03 | Opioid type dependence, in remission | F11.121 | Opioid abuse with intoxication, delirium |
| 304.1 | Sedative, hypnotic or anxiolytic dependence | F11.122 | Opioid abuse with intoxication, with perceptual disturbance |
| 304.10 | Sedative, hypnotic or anxiolytic dependence, unspecified | F11.129 | Opioid abuse with intoxication, unspecified |
| 304.11 | Sedative, hypnotic or anxiolytic dependence, continuous | F11.14 | Opioid abuse with opioid-induced mood disorder |
| 304.12 | Sedative, hypnotic or anxiolytic dependence, episodic | F11.15 | Opioid abuse with opioid-induced psychotic disorder |
| 304.13 | Sedative, hypnotic or anxiolytic dependence, in remission | F11.150 | Opioid abuse with opioid-induced psychotic disorder with delusions |
| 304.2 | Cocaine dependence | F11.151 | Opioid abuse with opioid-induced psychotic disorder with hallucinations |
| 304.20 | Cocaine dependence, unspecified | F11.159 | Opioid abuse with opioid-induced psychotic disorder, unspecified |
| 304.21 | Cocaine dependence, continuous | F11.18 | Opioid abuse with other opioid-induced disorder |
| 304.22 | Cocaine dependence, episodic | F11.181 | Opioid abuse with opioid-induced sexual dysfunction |
| 304.23 | Cocaine dependence, in remission | F11.182 | Opioid abuse with opioid-induced sleep disorder |
| 304.3 | Cannabis dependence | F11.188 | Opioid abuse with other opioid-induced disorder |
| 304.30 | Cannabis dependence, unspecified | F11.19 | Opioid abuse with unspecified opioid-induced disorder |
| 304.31 | Cannabis dependence, continuous | F11.2 | Opioid dependence |
| 304.32 | Cannabis dependence, episodic | F11.20 | Opioid dependence, uncomplicated |
| 304.33 | Cannabis dependence, in remission | F11.21 | Opioid dependence, in remission |

| | | | |
|---|---|---|---|
| 304.4 | Amphetamine and other psychostimulant dependence | F11.22 | Opioid dependence with intoxication |
| 304.40 | Amphetamine and other psychostimulant dependence, unspecified | F11.220 | Opioid dependence with intoxication, uncomplicated |
| 304.41 | Amphetamine and other psychostimulant dependence, continuous | F11.221 | Opioid dependence with intoxication delirium |
| 304.42 | Amphetamine and other psychostimulant dependence, episodic | F11.222 | Opioid dependence with intoxication with perceptual disturbance |
| 304.43 | Amphetamine and other psychostimulant dependence, in remission | F11.229 | Opioid dependence with intoxication, unspecified |
| 304.5 | Hallucinogen dependence | F11.23 | Opioid dependence with withdrawal |
| 304.50 | Hallucinogen dependence, unspecified | F11.24 | Opioid dependence with opioid-induced mood disorder |
| 304.51 | Hallucinogen dependence, continuous | F11.25 | Opioid dependence with opioid-induced psychotic disorder |
| 304.52 | Hallucinogen dependence, episodic | F11.250 | Opioid dependence with opioid-induced psychotic disorder with delusions |
| 304.53 | Hallucinogen dependence, in remission | F11.251 | Opioid dependence with opioid-induced psychotic disorder with hallucinations |
| 304.6 | Other specified drug dependence | F11.259 | Opioid dependence with opioid-induced psychotic disorder, unspecified |
| 304.60 | Other specified drug dependence, unspecified | F11.28 | Opioid dependence with other opioid-induced disorder |
| 304.61 | Other specified drug dependence, continuous | F11.281 | Opioid dependence with opioid-induced sexual dysfunction |
| 304.62 | Other specified drug dependence, episodic | F11.282 | Opioid dependence with opioid-induced sleep disorder |
| 304.63 | Other specified drug dependence, in remission | F11.288 | Opioid dependence with other opioid-induced disorder |
| 304.7 | Combinations of opioid type drug with any other drug dependence | F11.29 | Opioid dependence with unspecified opioid-induced disorder |
| 304.70 | Combinations of opioid type drug with any other drug dependence, unspecified | F12.1 | Cannabis abuse |
| 304.71 | Combinations of opioid type drug with any other drug dependence, continuous | F12.10 | Cannabis abuse, uncomplicated |
| 304.72 | Combinations of opioid type drug with any other drug dependence, episodic | F12.11 | Cannabis abuse, in remission |
| 304.73 | Combinations of opioid type drug with any other drug dependence, in remission | F12.12 | Cannabis abuse with intoxication |
| 304.8 | Combinations of drug dependence excluding opioid type drug | F12.120 | Cannabis abuse with intoxication |
| 304.80 | Combinations of drug dependence excluding opioid type drug, unspecified | F12.121 | Cannabis abuse with intoxication delirium |
| 304.81 | Combinations of drug dependence excluding opioid type drug, continuous | F12.122 | Cannabis abuse with intoxication with perceptual disturbance |
| 304.82 | Combinations of drug dependence excluding opioid type drug, episodic | F12.129 | Cannabis abuse with intoxication, unspecified |
| 304.83 | Combinations of drug dependence excluding opioid type drug, in remission | F12.15 | Cannabis abuse with psychotic disorder |
| 304.9 | Unspecified drug dependence | F12.150 | Cannabis abuse with psychotic disorder with delusions |
| 304.90 | Unspecified drug dependence, unspecified | F12.151 | Cannabis abuse with psychotic disorder with hallucinations |
| 304.91 | Unspecified drug dependence, continuous | F12.159 | Cannabis abuse with psychotic disorder, unspecified |
| 304.92 | Unspecified drug dependence, episodic | F12.18 | Cannabis abuse with other cannabis-induced disorder |

| | | | |
|---|---|---|---|
| 304.93 | Unspecified drug dependence, in remission | F12.180 | Cannabis abuse with cannabis-induced anxiety disorder |
| 305.2 | Nondependent cannabis abuse | F12.188 | Cannabis abuse with other cannabis-induced disorder |
| 305.20 | Cannabis abuse, unspecified | F12.19 | Cannabis abuse with unspecified cannabis-induced disorder |
| 305.21 | Cannabis abuse, continuous | F12.2 | Cannabis dependence |
| 305.22 | Cannabis abuse, episodic | F12.20 | Cannabis dependence, uncomplicated |
| 305.23 | Cannabis abuse, in remission | F12.21 | Cannabis dependence, in remission |
| 305.3 | Nondependent hallucinogen abuse | F12.22 | Cannabis dependence with intoxication |
| 305.30 | Hallucinogen abuse, unspecified | F12.220 | Cannabis dependence with intoxication, uncomplicated |
| 305.31 | Hallucinogen abuse, continuous | F12.221 | Cannabis dependence with intoxication delirium |
| 305.32 | Hallucinogen abuse, episodic | F12.222 | Cannabis dependence with intoxication with perceptual disturbance |
| 305.33 | Hallucinogen abuse, in remission | F12.229 | Cannabis dependence with intoxication, unspecified |
| 305.4 | Nondependent sedative, hypnotic or anxiolytic abuse | F12.23 | Cannabis dependence with withdrawal |
| 305.40 | Sedative, hypnotic or anxiolytic abuse, unspecified | F12.25 | Cannabis dependence with psychotic disorder |
| 305.41 | Sedative, hypnotic or anxiolytic abuse, continuous | F12.250 | Cannabis dependence with psychotic disorder with delusions |
| 305.42 | Sedative, hypnotic or anxiolytic abuse, episodic | F12.251 | Cannabis dependence with psychotic disorder with hallucinations |
| 305.43 | Sedative, hypnotic or anxiolytic abuse, in remission | F12.259 | Cannabis dependence with psychotic disorder, unspecified |
| 305.5 | Nondependent opioid abuse | F12.28 | Cannabis dependence with other cannabis-induced disorder |
| 305.50 | Opioid abuse, unspecified | F12.280 | Cannabis dependence with cannabis-induced anxiety disorder |
| 305.51 | Opioid abuse, continuous | F12.288 | Cannabis dependence with other cannabis-induced disorder |
| 305.52 | Opioid abuse, episodic | F12.29 | Cannabis dependence with unspecified cannabis-induced disorder |
| 305.53 | Opioid abuse, in remission | F12.90 | Cannabis use, uncomplicated |
| 305.6 | Nondependent cocaine abuse | F13.1 | Sedative, hypnotic or anxiolytic-related abuse |
| 305.60 | Cocaine abuse, unspecified | F13.10 | Sedative, hypnotic or anxiolytic abuse, uncomplicated |
| 305.61 | Cocaine abuse, continuous | F13.11 | Sedative, hypnotic or anxiolytic abuse, in remission |
| 305.62 | Cocaine abuse, episodic | F13.12 | Sedative, hypnotic or anxiolytic abuse with intoxication |
| 305.63 | Cocaine abuse, in remission | F13.120 | Sedative, hypnotic or anxiolytic abuse with intoxication, uncomplicated |
| 305.7 | Nondependent amphetamine or related acting sympathomimetic abuse | F13.121 | Sedative, hypnotic or anxiolytic abuse with intoxication delirium |
| 305.70 | Amphetamine or related acting sympathomimetic abuse, unspecified | F13.129 | Sedative, hypnotic or anxiolytic abuse with intoxication, unspecified |
| 305.71 | Amphetamine or related acting sympathomimetic abuse, continuous | F13.14 | Sedative, hypnotic or anxiolytic abuse with sedative, hypnotic or anxiolytic-induced mood disorder |

| | | | |
|---|---|---|---|
| 305.72 | Amphetamine or related acting sympathomimetic abuse, episodic | F13.15 | Sedative, hypnotic or anxiolytic abuse with sedative, hypnotic or anxiolytic-induced psychotic disorder |
| 305.73 | Amphetamine or related acting sympathomimetic abuse, in remission | F13.150 | Sedative, hypnotic or anxiolytic abuse with sedative, hypnotic or anxiolytic-induced psychotic disorder with delusions |
| 305.8 | Nondependent antidepressant type abuse | F13.151 | Sedative, hypnotic or anxiolytic abuse with sedative, hypnotic or anxiolytic-induced psychotic disorder with hallucinations |
| 305.80 | Antidepressant type abuse, unspecified | F13.159 | Sedative, hypnotic or anxiolytic abuse with sedative, hypnotic or anxiolytic-induced psychotic disorder, unspecified |
| 305.81 | Antidepressant type abuse, continuous | F13.18 | Sedative, hypnotic or anxiolytic abuse with other sedative, hypnotic or anxiolytic-induced disorders |
| 305.82 | Antidepressant type abuse, episodic | F13.180 | Sedative, hypnotic or anxiolytic abuse with sedative, hypnotic or anxiolytic-induced anxiety disorder |
| 305.83 | Antidepressant type abuse, in remission | F13.181 | Sedative, hypnotic or anxiolytic abuse with sedative, hypnotic or anxiolytic-induced sexual dysfunction |
| 305.9 | Nondependent other mixed or unspecified drug abuse | F13.182 | Sedative, hypnotic or anxiolytic abuse with sedative, hypnotic or anxiolytic-induced sleep disorder |
| 305.90 | Other, mixed, or unspecified drug abuse, unspecified | F13.188 | Sedative, hypnotic or anxiolytic abuse with other sedative, hypnotic or anxiolytic-induced disorder |
| 305.91 | Other, mixed, or unspecified drug abuse, continuous | F13.19 | Sedative, hypnotic or anxiolytic abuse with unspecified sedative, hypnotic or anxiolytic-induced disorder |
| 305.92 | Other, mixed, or unspecified drug abuse, episodic | F13.2 | Sedative, hypnotic or anxiolytic-related dependence |
| 305.93 | Other, mixed, or unspecified drug abuse, in remission | F13.20 | Sedative, hypnotic or anxiolytic dependence, uncomplicated |
| [redacted] | [redacted] | F13.21 | Sedative, hypnotic or anxiolytic dependence, in remission |
| [redacted] | [redacted] | F13.22 | Sedative, hypnotic or anxiolytic dependence with intoxication |
| [redacted] | [redacted] | F13.220 | Sedative, hypnotic or anxiolytic dependence with intoxication, uncomplicated |
| [redacted] | [redacted] | F13.221 | Sedative, hypnotic or anxiolytic dependence with intoxication delirium |
| [redacted] | [redacted] | F13.229 | Sedative, hypnotic or anxiolytic dependence with intoxication, unspecified |
| [redacted] | [redacted] | F13.23 | Sedative, hypnotic or anxiolytic dependence with withdrawal |
| [redacted] | [redacted] | F13.230 | Sedative, hypnotic or anxiolytic dependence with withdrawal, uncomplicated |
| [redacted] | [redacted] | F13.231 | Sedative, hypnotic or anxiolytic dependence with withdrawal delirium |
| [redacted] | [redacted] | F13.232 | Sedative, hypnotic or anxiolytic dependence with withdrawal with perceptual disturbance |
| [redacted] | [redacted] | F13.239 | Sedative, hypnotic or anxiolytic dependence with withdrawal, unspecified |
| [redacted] | [redacted] | F13.24 | Sedative, hypnotic or anxiolytic dependence with sedative, hypnotic or anxiolytic-induced mood disorder |
| [redacted] | [redacted] | F13.25 | Sedative, hypnotic or anxiolytic dependence with sedative, hypnotic or anxiolytic-induced psychotic disorder |
| [redacted] | [redacted] | F13.250 | Sedative, hypnotic or anxiolytic dependence with sedative, hypnotic or anxiolytic-induced psychotic disorder with delusions |

| | | | |
|---|---|---|---|
| [redacted] | | F13.251 | Sedative, hypnotic or anxiolytic dependence with sedative, hypnotic or anxiolytic-induced psychotic disorder with hallucinations |
| | | F13.259 | Sedative, hypnotic or anxiolytic dependence with sedative, hypnotic or anxiolytic-induced psychotic disorder, unspecified |
| | | F13.26 | Sedative, hypnotic or anxiolytic dependence with sedative, hypnotic or anxiolytic-induced persisting amnestic disorder |
| | | F13.27 | Sedative, hypnotic or anxiolytic dependence with sedative, hypnotic or anxiolytic-induced persisting dementia |
| | | F13.28 | Sedative, hypnotic or anxiolytic dependence with other sedative, hypnotic or anxiolytic-induced disorders |
| | | F13.280 | Sedative, hypnotic or anxiolytic dependence with sedative, hypnotic or anxiolytic-induced anxiety disorder |
| [redacted] | | F13.281 | Sedative, hypnotic or anxiolytic dependence with sedative, hypnotic or anxiolytic-induced sexual dysfunction |
| | | F13.282 | Sedative, hypnotic or anxiolytic dependence with sedative, hypnotic or anxiolytic-induced sleep disorder |
| | | F13.288 | Sedative, hypnotic or anxiolytic dependence with other sedative, hypnotic or anxiolytic-induced disorder |
| | | F13.29 | Sedative, hypnotic or anxiolytic dependence with unspecified sedative, hypnotic or anxiolytic-induced disorder |
| | | F14.1 | Cocaine abuse |
| | | F14.10 | Cocaine abuse, uncomplicated |
| | | F14.11 | Cocaine abuse, in remission |
| | | F14.12 | Cocaine abuse with intoxication |
| | | F14.120 | Cocaine abuse with intoxication, uncomplicated |
| | | F14.121 | Cocaine abuse with intoxication with delirium |
| | | F14.122 | Cocaine abuse with intoxication with perceptual disturbance |
| | | F14.129 | Cocaine abuse with intoxication, unspecified |
| | | F14.14 | Cocaine abuse with cocaine-induced mood disorder |
| [redacted] | | F14.15 | Cocaine abuse with cocaine-induced psychotic disorder |
| | | F14.150 | Cocaine abuse with cocaine-induced psychotic disorder with delusions |
| | | F14.151 | Cocaine abuse with cocaine-induced psychotic disorder with hallucinations |
| | | F14.159 | Cocaine abuse with cocaine-induced psychotic disorder, unspecified |
| | | F14.18 | Cocaine abuse with other cocaine-induced disorder |
| | | F14.180 | Cocaine abuse with cocaine-induced anxiety disorder |
| | | F14.181 | Cocaine abuse with cocaine-induced sexual dysfunction |

| | | | |
|---|---|---|---|
| [redacted] | | F14.182 | Cocaine abuse with cocaine-induced sleep disorder |
| | | F14.188 | Cocaine abuse with other cocaine-induced disorder |
| | | F14.19 | Cocaine abuse with unspecified cocaine-induced disorder |
| | | F14.2 | Cocaine dependence |
| | | F14.20 | Cocaine dependence, uncomplicated |
| | | F14.21 | Cocaine dependence, in remission |
| | | F14.22 | Cocaine dependence with intoxication |
| | | F14.220 | Cocaine dependence with intoxication, uncomplicated |
| | | F14.221 | Cocaine dependence with intoxication delirium |
| | | F14.222 | Cocaine dependence with intoxication with perceptual disturbance |
| | | F14.229 | Cocaine dependence with intoxication, unspecified |
| | | F14.23 | Cocaine dependence with withdrawal |
| | | F14.24 | Cocaine dependence with cocaine-induced mood disorder |
| | | F14.25 | Cocaine dependence with cocaine-induced psychotic disorder |
| | | F14.250 | Cocaine dependence with cocaine-induced psychotic disorder with delusions |
| | | F14.251 | Cocaine dependence with cocaine-induced psychotic disorder with hallucinations |
| | | F14.259 | Cocaine dependence with cocaine-induced psychotic disorder, unspecified |
| [redacted] | [redacted] | F14.28 | Cocaine dependence with other cocaine-induced disorder |
| | | F14.280 | Cocaine dependence with cocaine-induced anxiety disorder |
| | | F14.281 | Cocaine dependence with cocaine-induced sexual dysfunction |
| | | F14.282 | Cocaine dependence with cocaine-induced sleep disorder |
| | | F14.288 | Cocaine dependence with other cocaine-induced disorder |
| | | F14.29 | Cocaine dependence with unspecified cocaine-induced disorder |
| | | F15.1 | Other stimulant abuse |
| | | F15.10 | Other stimulant abuse, uncomplicated |
| | | F15.11 | Other stimulant abuse, in remission |
| | | F15.12 | Other stimulant abuse with intoxication |
| | | F15.120 | Other stimulant abuse with intoxication, uncomplicated |
| | | F15.121 | Other stimulant abuse with intoxication delirium |

| | | | |
|---|---|---|---|
| [redacted] | [redacted] | F15.122 | Other stimulant abuse with intoxication with perceptual disturbance |
| | | F15.129 | Other stimulant abuse with intoxication, unspecified |
| | | F15.14 | Other stimulant abuse with stimulant-induced mood disorder |
| | | F15.15 | Other stimulant abuse with stimulant-induced psychotic disorder |
| | | F15.150 | Other stimulant abuse with stimulant-induced psychotic disorder with delusions |
| | | F15.151 | Other stimulant abuse with stimulant-induced psychotic disorder with hallucinations |
| | | F15.159 | Other stimulant abuse with stimulant-induced psychotic disorder, unspecified |
| | | F15.18 | Other stimulant abuse with other stimulant-induced disorder |
| | | F15.180 | Other stimulant abuse with stimulant-induced anxiety disorder |
| | | F15.181 | Other stimulant abuse with stimulant-induced sexual dysfunction |
| | | F15.182 | Other stimulant abuse with stimulant-induced sleep disorder |
| | | F15.188 | Other stimulant abuse with other stimulant-induced disorder |
| | | F15.19 | Other stimulant abuse with unspecified stimulant-induced disorder |
| | | F15.2 | Other stimulant dependence |
| | | F15.20 | Other stimulant dependence, uncomplicated |
| | | F15.21 | Other stimulant dependence, in remission |
| | | F15.22 | Other stimulant dependence with intoxication |
| [redacted] | [redacted] | F15.220 | Other stimulant dependence with intoxication, uncomplicated |
| | | F15.221 | Other stimulant dependence with intoxication delirium |
| | | F15.222 | Other stimulant dependence with intoxication with perceptual disturbance |
| | | F15.229 | Other stimulant dependence with intoxication, unspecified |
| | | F15.23 | Other stimulant dependence with withdrawal |
| | | F15.24 | Other stimulant dependence with stimulant-induced mood disorder |
| | | F15.25 | Other stimulant dependence with stimulant-induced psychotic disorder |
| | | F15.250 | Other stimulant dependence with stimulant-induced psychotic disorder with delusions |
| | | F15.251 | Other stimulant dependence with stimulant-induced psychotic disorder with hallucinations |
| | | F15.259 | Other stimulant dependence with stimulant-induced psychotic disorder, unspecified |
| | | F15.28 | Other stimulant dependence with other stimulant-induced disorder |
| | | F15.280 | Other stimulant dependence with stimulant-induced anxiety disorder |

| | | |
|---|---|---|
| [redacted] | F15.281 | Other stimulant dependence with stimulant-induced sexual dysfunction |
| | F15.282 | Other stimulant dependence with stimulant-induced sleep disorder |
| | F15.288 | Other stimulant dependence with other stimulant-induced disorder |
| | F15.29 | Other stimulant dependence with unspecified stimulant-induced disorder |
| | F16.1 | Hallucinogen abuse |
| | F16.10 | Hallucinogen abuse, uncomplicated |
| | F16.11 | Hallucinogen abuse, in remission |
| | F16.12 | Hallucinogen abuse with intoxication |
| | F16.120 | Hallucinogen abuse with intoxication, uncomplicated |
| | F16.121 | Hallucinogen abuse with intoxication with delirium |
| | F16.122 | Hallucinogen abuse with intoxication with perceptual disturbance |
| | F16.129 | Hallucinogen abuse with intoxication, unspecified |
| | F16.14 | Hallucinogen abuse with hallucinogen-induced mood disorder |
| | F16.15 | Hallucinogen abuse with hallucinogen-induced psychotic disorder |
| | F16.150 | Hallucinogen abuse with hallucinogen-induced psychotic disorder with delusions |
| | F16.151 | Hallucinogen abuse with hallucinogen-induced psychotic disorder with hallucinations |
| | F16.159 | Hallucinogen abuse with hallucinogen-induced psychotic disorder, unspecified |
| | F16.18 | Hallucinogen abuse with other hallucinogen-induced disorder |
| | F16.180 | Hallucinogen abuse with hallucinogen-induced anxiety disorder |
| | F16.183 | Hallucinogen abuse with hallucinogen persisting perception disorder (flashbacks) |
| | F16.188 | Hallucinogen abuse with other hallucinogen-induced disorder |
| | F16.19 | Hallucinogen abuse with unspecified hallucinogen-induced disorder |
| | F16.2 | Hallucinogen dependence |
| | F16.20 | Hallucinogen dependence, uncomplicated |
| | F16.21 | Hallucinogen dependence, in remission |
| | F16.22 | Hallucinogen dependence with intoxication |
| | F16.220 | Hallucinogen dependence with intoxication, uncomplicated |
| | F16.221 | Hallucinogen dependence with intoxication with delirium |
| | F16.229 | Hallucinogen dependence with intoxication, unspecified |

| | | | |
|---|---|---|---|
| [redacted] | [redacted] | F16.24 | Hallucinogen dependence with hallucinogen-induced mood disorder |
| | | F16.25 | Hallucinogen dependence with hallucinogen-induced psychotic disorder |
| | | F16.250 | Hallucinogen dependence with hallucinogen-induced psychotic disorder with delusions |
| | | F16.251 | Hallucinogen dependence with hallucinogen-induced psychotic disorder with hallucinations |
| | | F16.259 | Hallucinogen dependence with hallucinogen-induced psychotic disorder, unspecified |
| | | F16.28 | Hallucinogen dependence with other hallucinogen-induced disorder |
| [redacted] | [redacted] | F16.280 | Hallucinogen dependence with hallucinogen-induced anxiety disorder |
| | | F16.283 | Hallucinogen dependence with hallucinogen persisting perception disorder (flashbacks) |
| | | F16.288 | Hallucinogen dependence with other hallucinogen-induced disorder |
| | | F16.29 | Hallucinogen dependence with unspecified hallucinogen-induced disorder |
| | | F18.1 | Inhalant abuse |
| | | F18.10 | Inhalant abuse, uncomplicated |
| | | F18.11 | Inhalant abuse, in remission |
| | | F18.12 | Inhalant abuse with intoxication |
| | | F18.120 | Inhalant abuse with intoxication, uncomplicated |
| | | F18.121 | Inhalant abuse with intoxication delirium |
| | | F18.129 | Inhalant abuse with intoxication, unspecified |
| | | F18.14 | Inhalant abuse with inhalant-induced mood disorder |
| | | F18.15 | Inhalant abuse with inhalant-induced psychotic disorder |
| | | F18.150 | Inhalant abuse with inhalant-induced psychotic disorder with delusions |
| | | F18.151 | Inhalant abuse with inhalant-induced psychotic disorder with hallucinations |
| | | F18.159 | Inhalant abuse with inhalant-induced psychotic disorder, unspecified |
| | | F18.17 | Inhalant abuse with inhalant-induced dementia |
| | | F18.18 | Inhalant abuse with other inhalant-induced disorders |
| | | F18.180 | Inhalant abuse with inhalant-induced anxiety disorder |
| | | F18.188 | Inhalant abuse with other inhalant-induced disorder |
| | | F18.19 | Inhalant abuse with unspecified inhalant-induced disorder |
| | | F18.2 | Inhalant dependence |
| | | F18.20 | Inhalant dependence, uncomplicated |

| | | |
|---|---|---|
| [redacted] | F18.21 | Inhalant dependence, in remission |
| | F18.22 | Inhalant dependence with intoxication |
| | F18.220 | Inhalant dependence with intoxication, uncomplicated |
| | F18.221 | Inhalant dependence with intoxication delirium |
| | F18.229 | Inhalant dependence with intoxication, unspecified |
| | F18.24 | Inhalant dependence with inhalant-induced mood disorder |
| | F18.25 | Inhalant dependence with inhalant-induced psychotic disorder |
| | F18.250 | Inhalant dependence with inhalant-induced psychotic disorder with delusions |
| | F18.251 | Inhalant dependence with inhalant-induced psychotic disorder with hallucinations |
| | F18.259 | Inhalant dependence with inhalant-induced psychotic disorder, unspecified |
| | F18.27 | Inhalant dependence with inhalant-induced dementia |
| | F18.28 | Inhalant dependence with other inhalant-induced disorders |
| | F18.280 | Inhalant dependence with inhalant-induced anxiety disorder |
| | F18.288 | Inhalant dependence with other inhalant-induced disorder |
| | F18.29 | Inhalant dependence with unspecified inhalant-induced disorder |
| | F19.1 | Other psychoactive substance abuse |
| | F19.10 | Other psychoactive substance abuse, uncomplicated |
| [redacted] | F19.11 | Other psychoactive substance abuse, in remission |
| | F19.12 | Other psychoactive substance abuse with intoxication |
| | F19.120 | Other psychoactive substance abuse with intoxication, uncomplicated |
| | F19.121 | Other psychoactive substance abuse with intoxication delirium |
| | F19.122 | Other psychoactive substance abuse with intoxication with perceptual disturbances |
| | F19.129 | Other psychoactive substance abuse with intoxication, unspecified |
| | F19.14 | Other psychoactive substance abuse with psychoactive substance-induced mood disorder |
| | F19.15 | Other psychoactive substance abuse with psychoactive substance-induced psychotic disorder |
| | F19.150 | Other psychoactive substance abuse with psychoactive substance-induced psychotic disorder with delusions |
| | F19.151 | Other psychoactive substance abuse with psychoactive substance-induced psychotic disorder with hallucinations |

| | | | |
|---|---|---|---|
| [redacted] | [redacted] | F19.159 | Other psychoactive substance abuse with psychoactive substance-induced psychotic disorder, unspecified |
| | | F19.16 | Other psychoactive substance abuse with psychoactive substance-induced persisting amnestic disorder |
| | | F19.17 | Other psychoactive substance abuse with psychoactive substance-induced persisting dementia |
| | | F19.18 | Other psychoactive substance abuse with other psychoactive substance-induced disorders |
| | | F19.180 | Other psychoactive substance abuse with psychoactive substance-induced anxiety disorder |
| | | F19.181 | Other psychoactive substance abuse with psychoactive substance-induced sexual dysfunction |
| [redacted] | [redacted] | F19.182 | Other psychoactive substance abuse with psychoactive substance-induced sleep disorder |
| | | F19.188 | Other psychoactive substance abuse with other psychoactive substance-induced disorder |
| | | F19.19 | Other psychoactive substance abuse with unspecified psychoactive substance-induced disorder |
| | | F19.2 | Other psychoactive substance dependence |
| | | F19.20 | Other psychoactive substance dependence, uncomplicated |
| | | F19.21 | Other psychoactive substance dependence, in remission |
| | | F19.22 | Other psychoactive substance dependence with intoxication |
| | | F19.220 | Other psychoactive substance dependence with intoxication, uncomplicated |
| | | F19.221 | Other psychoactive substance dependence with intoxication delirium |
| | | F19.222 | Other psychoactive substance dependence with intoxication with perceptual disturbance |
| | | F19.229 | Other psychoactive substance dependence with intoxication, unspecified |
| | | F19.23 | Other psychoactive substance dependence with withdrawal |
| | | F19.230 | Other psychoactive substance dependence with withdrawal, uncomplicated |
| [redacted] | [redacted] | F19.231 | Other psychoactive substance dependence with withdrawal delirium |
| | | F19.232 | Other psychoactive substance dependence with withdrawal with perceptual disturbance |
| | | F19.239 | Other psychoactive substance dependence with withdrawal, unspecified |
| | | F19.24 | Other psychoactive substance dependence with psychoactive substance-induced mood disorder |
| | | F19.25 | Other psychoactive substance dependence with psychoactive substance-induced psychotic disorder |
| | | F19.250 | Other psychoactive substance dependence with psychoactive substance-induced psychotic disorder with delusions |

| | | | |
|---|---|---|---|
| [redacted] | | F19.251 | Other psychoactive substance dependence with psychoactive substance-induced psychotic disorder with hallucinations |
| | | F19.259 | Other psychoactive substance dependence with psychoactive substance-induced psychotic disorder, unspecified |
| | | F19.26 | Other psychoactive substance dependence with psychoactive substance-induced persisting amnestic disorder |
| | | F19.27 | Other psychoactive substance dependence with psychoactive substance-induced persisting dementia |
| | | F19.28 | Other psychoactive substance dependence with other psychoactive substance-induced disorders |
| | | F19.280 | Other psychoactive substance dependence with psychoactive substance-induced anxiety disorder |
| [redacted] | | F19.281 | Other psychoactive substance dependence with psychoactive substance-induced sexual dysfunction |
| | | F19.282 | Other psychoactive substance dependence with psychoactive substance-induced sleep disorder |
| | | F19.288 | Other psychoactive substance dependence with other psychoactive substance-induced disorder |
| | | F19.29 | Other psychoactive substance dependence with unspecified psychoactive substance-induced disorder |
| **Mental Disorders** | | | |
| **ICD-9 Codes** | | **ICD-10 Codes** | |
| 295.0 | Simple type schizophrenia | F20 | Schizophrenia |
| 295.00 | Simple type schizophrenia, unspecified | F20.0 | Paranoid schizophrenia |
| 295.01 | Simple type schizophrenia, subchronic | F20.1 | Disorganized schizophrenia |
| 295.02 | Simple type schizophrenia, chronic | F20.2 | Catatonic schizophrenia |
| 295.03 | Simple type schizophrenia, subchronic with acute exacerbation | F20.3 | Undifferentiated schizophrenia |
| 295.04 | Simple type schizophrenia, chronic with acute exacerbation | F20.5 | Residual schizophrenia |
| 295.05 | Simple type schizophrenia, in remission | F20.8 | Other schizophrenia |
| 295.1 | Disorganized type schizophrenia | F20.81 | Schizophreniform disorder |
| 295.10 | Disorganized type schizophrenia, unspecified | F20.89 | Other schizophrenia |
| 295.11 | Disorganized type schizophrenia, subchronic | F20.9 | Schizophrenia, unspecified |
| 295.12 | Disorganized type schizophrenia, chronic | F25 | Schizoaffective disorders |
| 295.13 | Disorganized type schizophrenia, subchronic with acute exacerbation | F25.0 | Schizoaffective disorder, bipolar type |
| 295.14 | Disorganized type schizophrenia, chronic with acute exacerbation | F25.1 | Schizoaffective disorder, depressive type |
| 295.15 | Disorganized type schizophrenia, in remission | F25.8 | Other schizoaffective disorders |

| | | | |
|---|---|---|---|
| 295.2 | Catatonic type schizophrenia | F25.9 | Schizoaffective disorder, unspecified |
| 295.20 | Catatonic type schizophrenia, unspecified | F31 | Bipolar disorder |
| 295.21 | Catatonic type schizophrenia, subchronic | F31.0 | Bipolar disorder, current episode hypomanic |
| 295.22 | Catatonic type schizophrenia, chronic | F31.1 | Bipolar disorder, current episode manic without psychotic features |
| 295.23 | Catatonic type schizophrenia, subchronic with acute exacerbation | F31.10 | Bipolar disorder, current episode manic without psychotic features, unspecified |
| 295.24 | Catatonic type schizophrenia, chronic with acute exacerbation | F31.11 | Bipolar disorder, current episode manic without psychotic features, mild |
| 295.25 | Catatonic type schizophrenia, in remission | F31.12 | Bipolar disorder, current episode manic without psychotic features, moderate |
| 295.3 | Paranoid type schizophrenia | F31.13 | Bipolar disorder, current episode manic without psychotic features, severe |
| 295.30 | Paranoid type schizophrenia, unspecified | F31.2 | Bipolar disorder, current episode manic severe with psychotic features |
| 295.31 | Paranoid type schizophrenia, subchronic | F31.3 | Bipolar disorder, current episode depressed, mild or moderate severity |
| 295.32 | Paranoid type schizophrenia, chronic | F31.30 | Bipolar disorder, current episode depressed, mild or moderate severity, unspecified |
| 295.33 | Paranoid type schizophrenia, subchronic with acute exacerbation | F31.31 | Bipolar disorder, current episode depressed, mild |
| 295.34 | Paranoid type schizophrenia, chronic with acute exacerbation | F31.32 | Bipolar disorder, current episode depressed, moderate |
| 295.35 | Paranoid type schizophrenia, in remission | F31.4 | Bipolar disorder, current episode depressed, severe, without psychotic features |
| 295.4 | Schizophreniform disorder | F31.5 | Bipolar disorder, current episode depressed, severe, with psychotic features |
| 295.40 | Schizophreniform disorder, unspecified | F31.6 | Bipolar disorder, current episode mixed |
| 295.41 | Schizophreniform disorder, subchronic | F31.60 | Bipolar disorder, current episode mixed, unspecified |
| 295.42 | Schizophreniform disorder, chronic | F31.61 | Bipolar disorder, current episode mixed, mild |
| 295.43 | Schizophreniform disorder, subchronic with acute exacerbation | F31.62 | Bipolar disorder, current episode mixed, moderate |
| 295.44 | Schizophreniform disorder, chronic with acute exacerbation | F31.63 | Bipolar disorder, current episode mixed, severe, without psychotic features |
| 295.45 | Schizophreniform disorder, in remission | F31.64 | Bipolar disorder, current episode mixed, severe, with psychotic features |
| 295.5 | Latent schizophrenia | F31.7 | Bipolar disorder, currently in remission |
| 295.50 | Latent schizophrenia, unspecified | F31.70 | Bipolar disorder, currently in remission, most recent episode unspecified |
| 295.51 | Latent schizophrenia, subchronic | F31.71 | Bipolar disorder, in partial remission, most recent episode hypomanic |
| 295.52 | Latent schizophrenia, chronic | F31.72 | Bipolar disorder, in full remission, most recent episode hypomanic |
| 295.53 | Latent schizophrenia, subchronic with acute exacerbation | F31.73 | Bipolar disorder, in partial remission, most recent episode manic |
| 295.54 | Latent schizophrenia, chronic with acute exacerbation | F31.74 | Bipolar disorder, in full remission, most recent episode manic |
| 295.55 | Latent schizophrenia, in remission | F31.75 | Bipolar disorder, in partial remission, most recent episode depressed |
| 295.6 | Schizophrenic disorder, residual type | F31.76 | Bipolar disorder, in full remission, most recent episode depressed |

| | | | |
|---|---|---|---|
| 295.60 | Schizophrenic disorders, residual type, unspecified | F31.77 | Bipolar disorder, in partial remission, most recent episode mixed |
| 295.61 | Schizophrenic disorders, residual type, subchronic | F31.78 | Bipolar disorder, in full remission, most recent episode mixed |
| 295.62 | Schizophrenic disorders, residual type, chronic | F31.8 | Other bipolar disorders |
| 295.63 | Schizophrenic disorders, residual type, subchronic with acute exacerbation | F31.81 | Bipolar II disorder |
| 295.64 | Schizophrenic disorders, residual type, chronic with acute exacerbation | F31.89 | Other bipolar disorder |
| 295.65 | Schizophrenic disorders, residual type, in remission | F31.9 | Bipolar disorder, unspecified |
| 295.8 | Other specified types of schizophrenia | F30.10 | Manic episode without psychotic symptoms, unspecified |
| 295.80 | Other specified types of schizophrenia, unspecified | F30.11 | Manic episode without psychotic symptoms, mild |
| 295.81 | Other specified types of schizophrenia, subchronic | F30.12 | Manic episode without psychotic symptoms, moderate |
| 295.82 | Other specified types of schizophrenia, chronic | F30.13 | Manic episode, severe, without psychotic symptoms |
| 295.83 | Other specified types of schizophrenia, subchronic with acute exacerbation | F30.2 | Manic episode, severe with psychotic symptoms |
| 295.84 | Other specified types of schizophrenia, chronic with acute exacerbation | F30.3 | Manic episode in partial remission |
| 295.85 | Other specified types of schizophrenia, in remission | F30.4 | Manic episode in full remission |
| 295.9 | Unspecified schizophrenia | F30.8 | Other manic episodes |
| 295.90 | Unspecified schizophrenia, unspecified | F32.89 | Other specified depressive episodes |
| 295.91 | Unspecified schizophrenia, subchronic | F39 | Unspecified mood [affective] disorder |
| 295.92 | Unspecified schizophrenia, chronic | F34.81 | Disruptive mood dysregulation disorder |
| 295.93 | Unspecified schizophrenia, subchronic with acute exacerbation | F34.89 | Other specified persistent mood disorders |
| 295.94 | Unspecified schizophrenia, chronic with acute exacerbation | F32.9 | Major depressive disorder, single episode, unspecified |
| 295.95 | Unspecified schizophrenia, in remission | F32.0 | Major depressive disorder, single episode, mild |
| V11.0 | Schizoaffective disorder | F32.1 | Major depressive disorder, single episode, moderate |
| 295.7 | Schizoaffective disorder | F32.2 | Major depressive disorder, single episode, severe without psychotic features |
| 295.70 | Schizoaffective disorder, unspecified | F32.3 | Major depressive disorder, single episode, severe with psychotic features |
| 295.71 | Schizoaffective disorder, subchronic | F32.4 | Major depressive disorder, single episode, in partial remission |
| 295.72 | Schizoaffective disorder, chronic | F32.5 | Major depressive disorder, single episode, in full remission |
| 295.73 | Schizoaffective disorder, subchronic with acute exacerbation | F33.0 | Major depressive disorder, recurrent, mild |
| 295.74 | Schizoaffective disorder, chronic with acute exacerbation | F33.1 | Major depressive disorder, recurrent, moderate |
| 295.75 | Schizoaffective disorder, in remission | F33.2 | Major depressive disorder, recurrent severe without psychotic features |
| 296.0 | Bipolar I disorder, single manic episode | F33.3 | Major depressive disorder, recurrent, severe with psychotic symptoms |

| | | | |
|---|---|---|---|
| 296.00 | Bipolar I disorder, single manic episode, unspecified | F33.4 | Major depressive disorder, recurrent, in remission |
| 296.01 | Bipolar I disorder, single manic episode, mild | F33.40 | Major depressive disorder, recurrent, in remission, unspecified |
| 296.02 | Bipolar I disorder, single manic episode, moderate | F33.41 | Major depressive disorder, recurrent, in partial remission |
| 296.03 | Bipolar I disorder, single manic episode, severe, without mention of psychotic behavior | F33.42 | Major depressive disorder, recurrent, in full remission |
| 296.04 | Bipolar I disorder, single manic episode, severe, specified as with psychotic behavior | F33.9 | Major depressive disorder, recurrent, unspecified |
| 296.05 | Bipolar I disorder, single manic episode, in partial or unspecified remission | F41 | Other anxiety disorders |
| 296.06 | Bipolar I disorder, single manic episode, in full remission | F41.0 | Panic disorder [episodic paroxysmal anxiety] |
| 296.1 | Manic disorder recurrent episode | F41.1 | Generalized anxiety disorder |
| 296.10 | Manic affective disorder, recurrent episode, unspecified | F41.3 | Other mixed anxiety disorders |
| 296.11 | Manic affective disorder, recurrent episode, mild | F41.8 | Other specified anxiety disorders |
| 296.12 | Manic affective disorder, recurrent episode, moderate | F41.9 | Anxiety disorder, unspecified |
| 296.13 | Manic affective disorder, recurrent episode, severe, without mention of psychotic behavior | F43.1 | Post-traumatic stress disorder (PTSD) |
| 296.14 | Manic affective disorder, recurrent episode, severe, specified as with psychotic behavior | F43.10 | Post-traumatic stress disorder, unspecified |
| 296.15 | Manic affective disorder, recurrent episode, in partial or unspecified remission | F43.11 | Post-traumatic stress disorder, acute |
| 296.16 | Manic affective disorder, recurrent episode, in full remission | F43.12 | Post-traumatic stress disorder, chronic |
| 296.4 | Bipolar I disorder, most recent episode (or current) manic | [redacted] | |
| 296.40 | Bipolar I disorder, most recent episode (or current) manic, unspecified | | |
| 296.41 | Bipolar I disorder, most recent episode (or current) manic, mild | [redacted] | |
| 296.42 | Bipolar I disorder, most recent episode (or current) manic, moderate | | |
| 296.43 | Bipolar I disorder, most recent episode (or current) manic, severe, without mention of psychotic behavior | | |
| 296.44 | Bipolar I disorder, most recent episode (or current) manic, severe, specified as with psychotic behavior | | |
| 296.45 | Bipolar I disorder, most recent episode (or current) manic, in partial or unspecified remission | | |
| 296.46 | Bipolar I disorder, most recent episode (or current) manic, in full remission | | |
| 296.5 | Bipolar I disorder, most recent episode (or current) depressed | | |
| 296.50 | Bipolar I disorder, most recent episode (or current) depressed, unspecified | | |
| 296.51 | Bipolar I disorder, most recent episode (or current) depressed, mild | | |
| 296.52 | Bipolar I disorder, most recent episode (or current) depressed, moderate | | |
| 296.53 | Bipolar I disorder, most recent episode (or current) depressed, severe, without mention of psychotic behavior | | |
| 296.54 | Bipolar I disorder, most recent episode (or current) depressed, severe, specified as with psychotic behavior | | |

| | | | |
|---|---|---|---|
| 296.55 | Bipolar I disorder, most recent episode (or current) depressed, in partial or unspecified remission | [redacted] | [redacted] |
| 296.56 | Bipolar I disorder, most recent episode (or current) depressed, in full remission | [redacted] | [redacted] |
| 296.6 | Bipolar I disorder, most recent episode (or current) mixed | [redacted] | [redacted] |
| 296.60 | Bipolar I disorder, most recent episode (or current) mixed, unspecified | [redacted] | [redacted] |
| 296.61 | Bipolar I disorder, most recent episode (or current) mixed, mild | [redacted] | [redacted] |
| 296.62 | Bipolar I disorder, most recent episode (or current) mixed, moderate | [redacted] | [redacted] |
| 296.63 | Bipolar I disorder, most recent episode (or current) mixed, severe, without mention of psychotic behavior | [redacted] | [redacted] |
| 296.64 | Bipolar I disorder, most recent episode (or current) mixed, severe, specified as with psychotic behavior | [redacted] | [redacted] |
| 296.65 | Bipolar I disorder, most recent episode (or current) mixed, in partial or unspecified remission | [redacted] | [redacted] |
| 296.66 | Bipolar I disorder, most recent episode (or current) mixed, in full remission | [redacted] | [redacted] |
| 296.7 | Bipolar I disorder, most recent episode (or current) unspecified | [redacted] | [redacted] |
| 296.8 | Other and unspecified bipolar disorders | [redacted] | [redacted] |
| 296.80 | Bipolar disorder, unspecified | [redacted] | [redacted] |
| 296.81 | Atypical manic disorder | [redacted] | [redacted] |
| 296.82 | Atypical depressive disorder | [redacted] | [redacted] |
| 296.89 | Other bipolar disorders | [redacted] | [redacted] |
| 296.9 | Other and unspecified episodic mood disorder | [redacted] | [redacted] |
| 296.90 | Unspecified episodic mood disorder | [redacted] | [redacted] |
| 296.99 | Other specified episodic mood disorder | [redacted] | [redacted] |
| V11.1 | Personal history of affective disorders | [redacted] | [redacted] |
| 296.20 | Major depressive affective disorder, single episode, unspecified | [redacted] | [redacted] |
| 296.21 | Major depressive affective disorder, single episode, mild | [redacted] | [redacted] |
| 296.22 | Major depressive affective disorder, single episode, moderate | [redacted] | [redacted] |
| 296.23 | Major depressive affective disorder, single episode, severe, without mention of psychotic behavior | [redacted] | [redacted] |
| 296.24 | Major depressive affective disorder, single episode, severe, specified as with psychotic behavior | [redacted] | [redacted] |
| 296.25 | Major depressive affective disorder, single episode, in partial or unspecified remission | [redacted] | [redacted] |
| 296.26 | Major depressive affective disorder, single episode, in full remission | [redacted] | [redacted] |
| 296.3 | Major depressive disorder recurrent episode | [redacted] | [redacted] |
| 296.30 | Major depressive affective disorder, recurrent episode, unspecified | [redacted] | [redacted] |

| | | |
|---|---|---|
| 296.31 | Major depressive affective disorder, recurrent episode, mild | [redacted] |
| 296.32 | Major depressive affective disorder, recurrent episode, moderate | |
| 296.33 | Major depressive affective disorder, recurrent episode, severe, without mention of psychotic behavior | |
| 296.34 | Major depressive affective disorder, recurrent episode, severe, specified as with psychotic behavior | |
| 296.35 | Major depressive affective disorder, recurrent episode, in partial or unspecified remission | |
| 296.36 | Major depressive affective disorder, recurrent episode, in full remission | |
| 300.00 | Anxiety state, unspecified | |
| 300.01 | Panic disorder without agoraphobia | |
| 300.02 | Generalized anxiety disorder | |
| 300.09 | Other anxiety states | |
| 799.2 | Nervousness | |
| 309.81 | Posttraumatic stress disorder | |

Note: If the codes were given in the outpatient setting, two codes were required for a diagnosis. If the codes were given in the inpatient setting, only one code was required for a diagnosis.

Supplemental Figure 3a. Percentage of men with an AUD diagnosis by maximum binge drinking frequency, stratified by race/ethnicity (n=638,166).

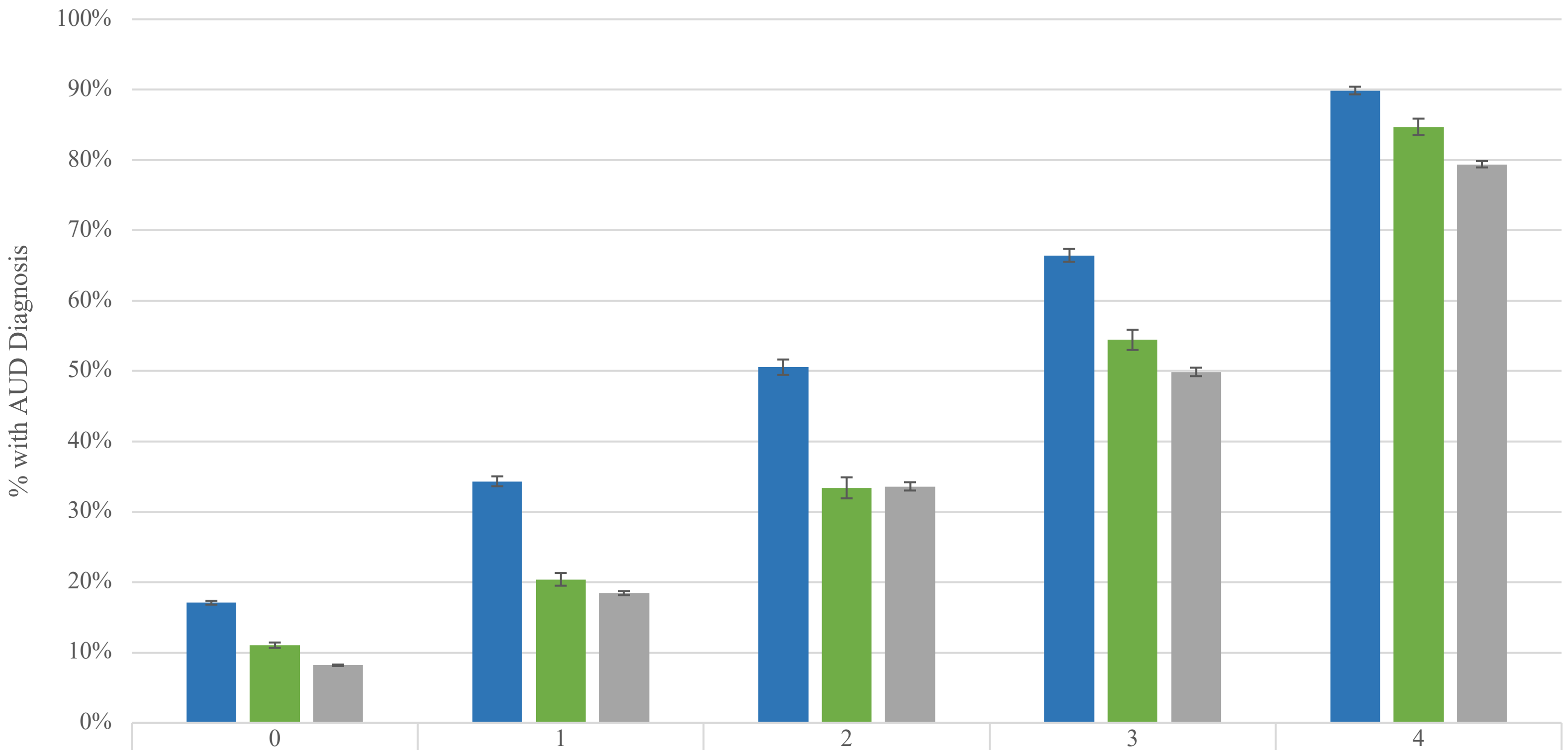


| | 0 | 1 | 2 | 3 | 4 |
|---|---|---|---|---|---|
| Black | 17.1% | 34.3% | 50.6% | 66.5% | 89.9% |
| Hispanic | 11.1% | 20.4% | 33.4% | 54.5% | 84.7% |
| White | 8.2% | 18.4% | 33.6% | 49.9% | 79.4% |

Binge Drinking Frequency

AUD=alcohol use disorder; AUDIT-C=Alcohol Use Disorders Identification Test – Consumption.

Note: Binge drinking frequency is measured by the 3rd item of the AUDIT-C where 0=never, 1=less than monthly, 2=monthly, 3=weekly, and 4=daily or almost daily.

Supplemental Figure 3b. Percentage of women with an AUD diagnosis by maximum binge drinking frequency, stratified by race/ethnicity (n=61,805).

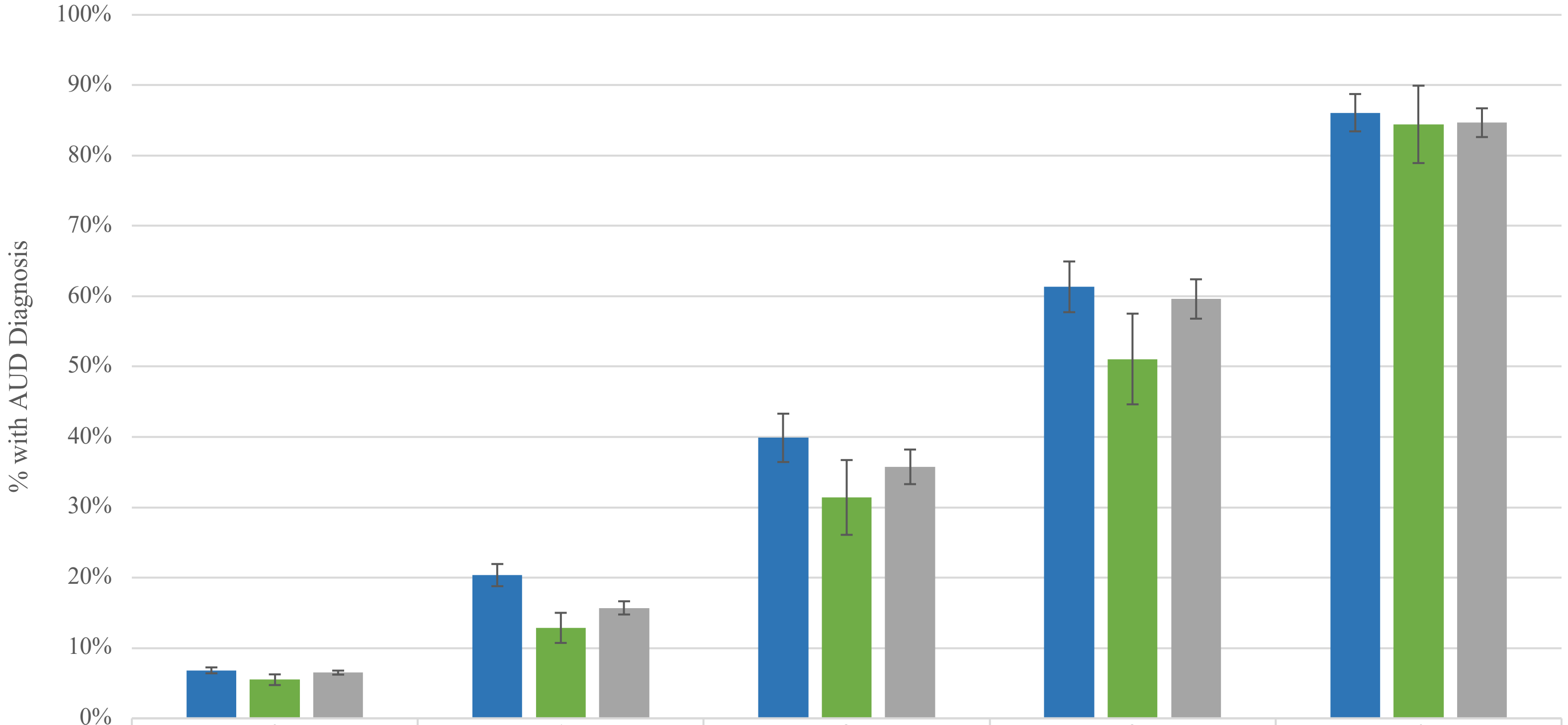


| | 0 | 1 | 2 | 3 | 4 |
|---|---|---|---|---|---|
| Black | 6.8% | 20.4% | 39.9% | 61.3% | 86.1% |
| Hispanic | 5.5% | 12.9% | 31.4% | 51.1% | 84.4% |
| White | 6.5% | 15.7% | 35.8% | 59.6% | 84.7% |

AUD=alcohol use disorder; AUDIT-C=Alcohol Use Disorders Identification Test – Consumption.
Note: Binge drinking frequency is measured by the 3rd item of the AUDIT-C where 0=never, 1=less than monthly, 2=monthly, 3=weekly, and 4=daily or almost daily.

Supplemental Figure 4. Predicted probabilities of AUD diagnosis among women, by race/ethnicity (n=61,803).

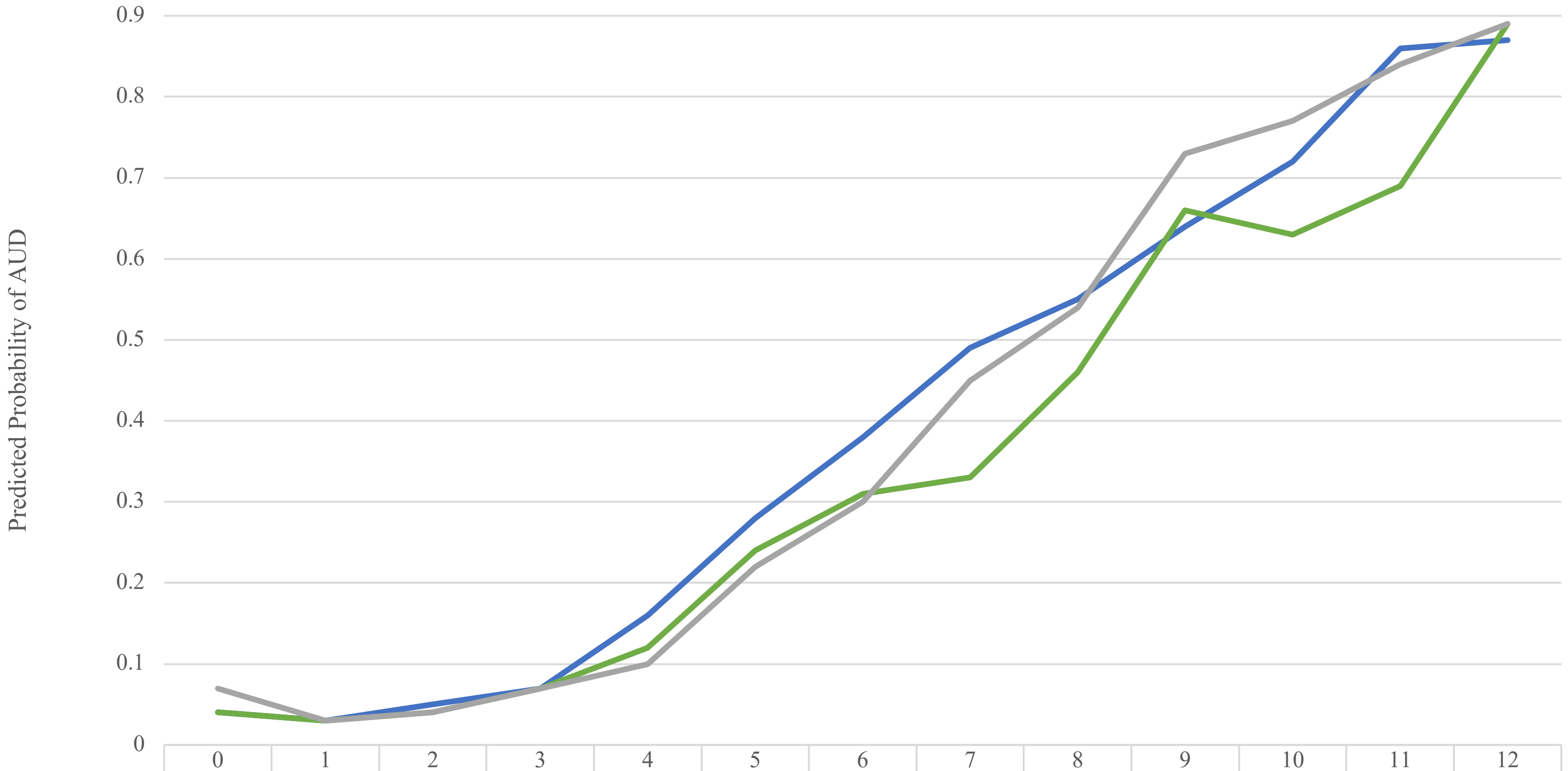


| | 0 | 1 | 2 | 3 | 4 | 5 | 6 | 7 | 8 | 9 | 10 | 11 | 12 |
|---|---|---|---|---|---|---|---|---|---|---|---|---|---|
| Black | 0.04 | 0.03 | 0.05 | 0.07 | 0.16 | 0.28 | 0.38 | 0.49 | 0.55 | 0.64 | 0.72 | 0.86 | 0.87 |
| Hispanic | 0.04 | 0.03 | 0.04 | 0.07 | 0.12 | 0.24 | 0.31 | 0.33 | 0.46 | 0.66 | 0.63 | 0.69 | 0.89 |
| White | 0.07 | 0.03 | 0.04 | 0.07 | 0.10 | 0.22 | 0.30 | 0.45 | 0.54 | 0.73 | 0.77 | 0.84 | 0.89 |

Maximum AUDIT-C Score

AUD=alcohol use disorder; AUDIT-C= Alcohol Use Disorders Identification Test – Consumption.

Note: These estimates come from the multivariable logistic regression model of factors associated with AUD diagnosis (Table 2). Age was set at 51 years (mean age for women), mental disorder=1, all other covariates=0. The connecting lines are for visualization purposes only since the interaction was accounted for by a composite variable of race/ethnicity and maximum AUDIT-C score, and should not be interpreted as continuous.

Supplemental Table 3. Sensitivity analysis of factors associated with AUD diagnosis using age-adjusted mean AUDIT-C score as the measure of alcohol consumption, overall and stratified by sex.

| | Overall (n=699,953)* | | Men (n=638,150)* | | Women (n=61,803)* | |
|---|---|---|---|---|---|---|
| *Variables* | aOR (95% CI) | *P* | aOR (95% CI) | *P* | aOR (95% CI) | *P* |
| Race/ethnicity x age-adjusted mean AUDIT-C score** | | | | | | |
| Black (ref=White) | 2.15 (2.04, 2.26) | <0.0001 | 2.15 (2.04, 2.26) | <0.0001 | 1.67 (1.45, 1.92) | <0.0001 |
| Hispanic (ref=White) | 1.23 (1.14, 1.32) | <0.0001 | 1.21 (1.12, 1.31) | <0.0001 | 1.27 (1.02, 1.59) | 0.0329 |
| Women (ref=men) | 0.59 (0.57, 0.61) | <0.0001 | - | - | - | - |
| Age (10-yr increments) | 1.01 (1.01, 1.01) | <0.0001 | 1.01 (1.01, 1.01) | 0.0002 | 1.04 (1.04, 1.04) | 0.0023 |
| *Alcohol-Related Characteristics* | | | | | | |
| Cirrhosis*** | 20.02 (18.64, 21.49) | <0.0001 | 19.72 (18.35, 21.18) | <0.0001 | 26.21 (15.99, 42.97) | <0.0001 |
| Neuropathy*** | 21.19 (17.19, 26.14) | <0.0001 | 20.17 (16.34, 24.89) | <0.0001 | 319.13 (35.66, >999.99) | <0.0001 |
| Cardiomyopathy*** | 21.13 (17.16, 26.02) | <0.0001 | 20.79 (16.86, 25.62) | <0.0001 | 41.96 (6.22, 283.14) | 0.0001 |
| Gastritis*** | 46.10 (30.33, 70.05) | <0.0001 | 47.44 (30.78, 73.10) | <0.0001 | 22.98 (4.57, 115.62) | 0.0001 |
| Fatty liver disease*** | 5.52 (4.94, 6.17) | <0.0001 | 5.56 (4.96, 6.23) | <0.0001 | 4.95 (2.76, 8.90) | <0.0001 |
| Hepatitis*** | 21.15 (16.98, 26.35) | <0.0001 | 22.11 (17.64, 27.71) | <0.0001 | 6.86 (2.73, 17.27) | <0.0001 |
| Liver damage*** | 10.52 (8.60, 12.87) | <0.0001 | 10.10 (8.23, 12.40) | <0.0001 | 29.02 (9.61, 87.60) | <0.0001 |
| *Other Clinical ICD-9/10 Diagnoses* | | | | | | |
| Drug abuse/dependence | 14.77 (14.50, 15.05) | <0.0001 | 14.60 (14.32, 14.89) | <0.0001 | 15.75 (14.75, 16.82) | <0.0001 |
| Mental disorder | 3.96 (3.89, 4.03) | <0.0001 | 3.88 (3.81, 3.95) | <0.0001 | 6.96 (6.22, 7.80) | <0.0001 |
| C-statistic | 0.88 | | 0.88 | | 0.88 | |

aOR=adjusted odds ratio; 95% CI =95% confidence interval; AUDIT-C=Alcohol Use Disorders Identification Test – Consumption; AUD=alcohol use disorder. *Number of observations with missing age: Black men (n=5); Hispanic men (n=8); White men (n=42); Black women (n=1); Hispanic women (n=1); White women(n=3). **Odds ratios displayed at age-adjusted mean AUDIT-C scores equal to those in the primary analysis (mean=3, 3, 2 for overall, men, and women, respectively). ***Alcohol-specific diagnosis.
